# Expansion, persistence and efficacy of donor memory-like NK cells for the treatment of post-transplant relapse

**DOI:** 10.1101/2021.08.24.21262547

**Authors:** Roman M. Shapiro, Grace Birch, Juliana Vergara, Guangan Hu, Sarah Nikiforow, Joanna Baginska, Alaa Ali, Mubin Tarannum, Benedetta Rambaldi, Yohei Arihara, Carol Reynolds, Max Halpern, Scott Rodig, Nicole Cullen, Andrew A. Lane, R. Coleman Lindsley, Corey S. Cutler, Joseph H. Antin, Vincent T. Ho, John Koreth, Mahasweta Gooptu, Haesook T. Kim, Karl-Johan Malmberg, Catherine J. Wu, Jianzhu Chen, Robert J. Soiffer, Jerome Ritz, Rizwan Romee

## Abstract

**Background:** Responses to conventional donor lymphocyte infusion (DLI) for post-allogeneic hematopoietic cell transplantation (HCT) relapse are typically poor. Natural killer (NK) cell-based therapy is a promising modality to treat post-HCT relapse.

**Methods:** We initiated this ongoing phase 1 trial of adoptively transferred **c**ytokine ***i***nduced ***m***emory-***l***ike (CIML) NK cells in patients with myeloid malignancies relapsed after haploidentical HCT. All patients received a donor-derived NK cell dose of 5-10 × million cells/kg after lymphodepleting chemotherapy, followed by systemic IL-2 for 7 doses. High resolution profiling with mass cytometry and single cell RNA sequencing characterized the expanding and persistent NK cells subpopulations in a longitudinal manner after infusion. *In vitro* functional studies of infused CIML NK cells were complemented with *in vivo* evaluation of NK trafficking to disease sites using multiparameter immunofluorescence.

**Results:** In the first 5 patients on the trial, infusion of CIML NK cells led to a rapid 10 to 50-fold *in vivo* expansion that was sustained over months. The infusion was well-tolerated, with fever and pancytopenia as the most common adverse events. Responses were attained in 4 of 5 patients at day +28. Immunophenotypic and transcriptional profiling revealed a dynamic evolution of the activated CIML NK cell phenotype, superimposed on the natural variation in donor NK cell repertoires. AML relapse after initial response to CIML NK cell therapy was associated with low transcript expression of CD2 ligands, including *CD48/SLAMF2* and CD58/*LFA3*.

**Conclusion:** Given their rapid expansion and long-term persistence in an immune compatible environment, CIML NK cells serve as a promising platform for the treatment of post-transplant relapse of myeloid disease. Further characterization of their unique in vivo biology and interaction with both T-cells and tumor targets will lead to the development of novel cell-based immunotherapies.

## Introduction

Relapse of acute myeloid leukemia (AML) occurs in up to 50% of patients after allogeneic hematopoietic stem cell transplant (HCT) and heralds an extremely poor prognosis[1]. In several series of patients with AML who relapsed following HCT, the median overall survival was less than 6 months [2]. Traditional approaches for the treatment of post-HCT relapse include the use of donor lymphocyte infusions (DLI) or additional chemotherapy followed by a 2^nd^ HCT [1, 3]. While DLI is associated with remission rates in the 10-25% range, it is also associated with a high risk of moderate to severe graft versus host (GVHD), and long-term survival with this approach remains poor [4]. With the recent rise in haploidentical HCT (haplo-HCT) whereby it is less clear whether the risk of GVHD due to DLI is outweighed by its efficacy [5, 6], the treatment of post-transplant relapse must be improved.

The development of novel approaches for the management of post-HCT relapse of myeloid disease remains a high priority area of research. Early post-HCT reconstitution of natural killer (NK) cells has been associated with a reduced risk of relapse in a number of retrospective studies, including in the T-cell depleted setting [8-10]. NK cells are an attractive platform for the management of post-HCT relapse because they are capable of mediating a graft-versus-leukemia (GvL) effect without causing GVHD [10-13]. Evidence supporting a role in GvL emanates from studies demonstrating that mismatch in killer-cell Ig like receptor (KIR) antigens, including HLA class I molecules, between NK cells and their putative targets is involved in NK cell activation and cytotoxicity [6, 14]. While this is believed to be particularly important in haplo-HCT, the mediation of NK alloreactivity by KIR-independent recognition of target ligands through activating receptors on the surface of NK cells and modulation of these effects by T-cells in the stem cell graft suggest that NK cells may promote a substantial GvL effect in HLA-matched transplants as well [11].

The adoptive transfer of NK cells in both the pre- and post-transplant settings has had mixed results in prior clinical trials [11]. The heterogeneity in clinical efficacy of predicted NK cell alloreactivity has been attributed in part to non-persistence of NK cells following infusion, inhibition from T-regulatory or myeloid suppressor cells, and tumor-induced NK cell anergy [15, 16]. Strategies to overcome these barriers focus on generating a sufficient quantity of NK cells that persist *in vivo* and retain their efficacy against potential target cells [17-19].

The incubation of donor-derived NK cells with the cytokines IL-12, IL-15, IL-18 generates a phenotype known as cytokine-induced memory-like (CIML) NK cells, capable of persistent survival, expansion, and avoidance of anergy *in vivo* [20, 21]. In pre-clinical studies, CIML NK cells exhibited a lower threshold of activation in response to cytokine restimulation as well as enhanced cytotoxicity against target leukemia cells [21]. In a phase 1 trial of adoptively transferred CIML NK cell from a haploidentical donor in patients whose AML was relapsed/refractory and who never received a stem cell transplant, CR/CRi (complete remission with insufficient hematological recovery) was attained in greater than 50% of subjects [20, 22]. While exhibiting enhanced cytotoxicity against leukemia, the CIML NK cells only persisted for 2-4 weeks in the treated patients due to HLA incompatibility. In contrast, adoptive transfer of CIML NK cells in the context of a syngeneic mouse model resulted in persistence of functional cells for several months, arguing that their transfer into an immune compatible environment prolongs their survival [23].

The prolonged persistence of functional CIML NK cells in an immune compatible environment supports their use in the post-HCT setting, a setting in which immune compatibility is possible because of the common donor for both stem cells and subsequent adoptive transfer of NK cells. Coupled with the demonstrated anti-leukemia activity of CIML NK cells in the HLA-mismatched setting, we initiated a phase I trial of CIML NK cells to treat patients with relapsed myeloid malignancies after haplo-HCT. Here we describe the detailed characterization of the CIML NK cells *in vivo* in the first 5 patients treated on this trial, including transcriptome and proteome analysis at single cell resolution.

## Results

### Adoptive transfer of donor-derived CIML NK cells is safe and associated with clinical responses

Five patients whose myeloid disease relapsed after haplo-HCT were treated with a donor-derived CIML NK cell product (**Figure 1A**). The five patients included 3 with acute myeloid leukemia (AML), 1 with myelodysplastic syndrome (MDS), and 1 with blastic plasmacytoid dendritic cell neoplasm (BPDCN). Characteristics of the patients are shown in the Appendix (**Table S1**). As per trial inclusion criteria, none of the patients received any prior donor lymphocyte infusions (DLI), and none received any other therapy concurrent with the CIML NK cell infusion. Patients tolerated the CIML NK cell infusion and expansion well, with the most common side effect being fever (temperature ranging from 38.1°C to 39.3°C) during the 12 days of IL-2 administration. One patient developed grade 2 cytokine release syndrome (CRS) and was treated with tocilizumab. Four of the first five patients experienced pancytopenia. In two cases the pancytopenia was prolonged and two patients received CD34^+^ cell selected stem cell boosts from their original donors 5 and 6 weeks after CIML NK therapy, respectively. Both patients responded to the CD34^+^ cell selected stem cell boosts, with neutrophil recovery evident within 9-14 days post boost. No patient developed any evidence of GvHD.

**Figure 1.**
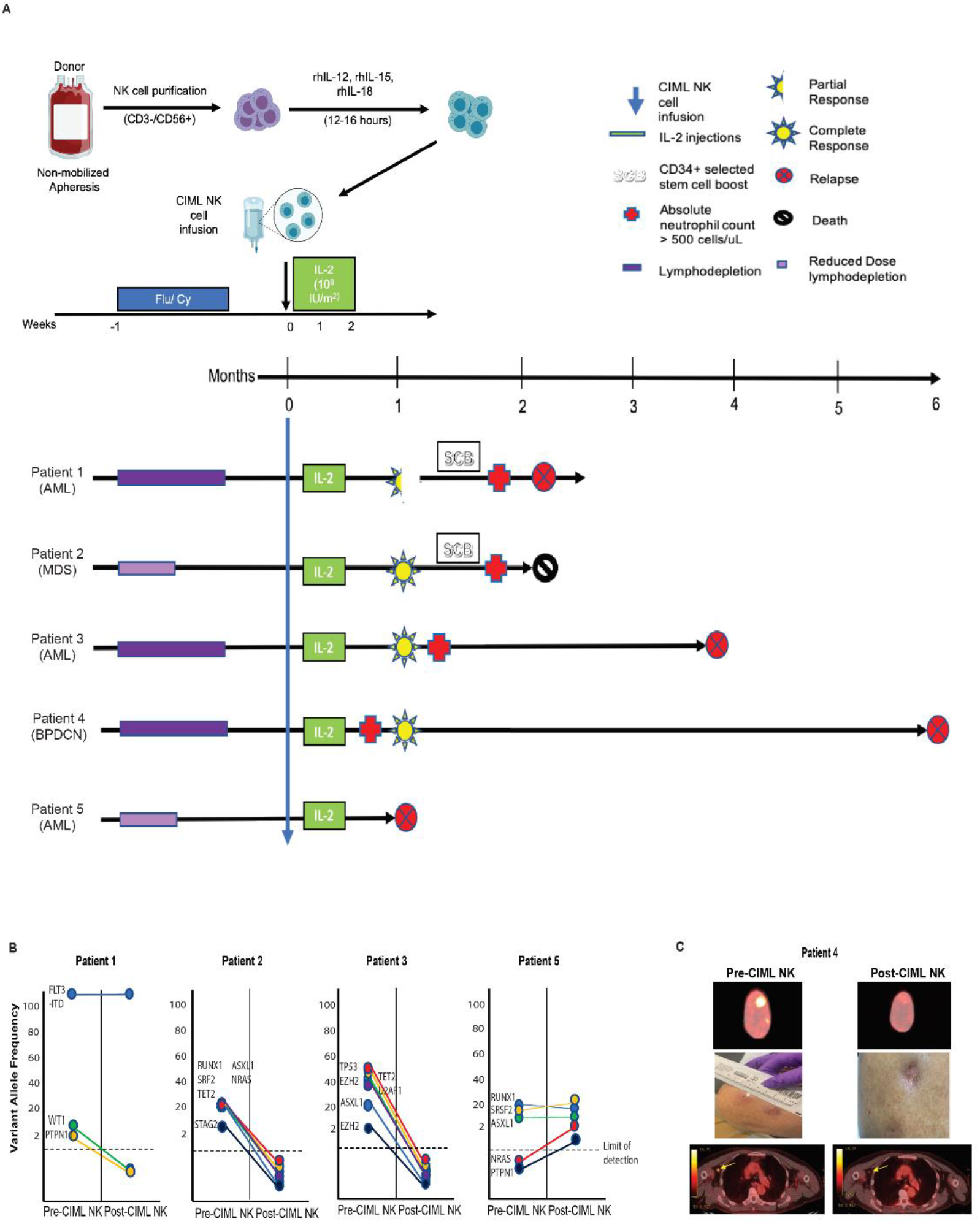
Phase I CIML NK cell trial overview and clinical responses. **A** Schema of the CIML NK trial therapy. For each patient, the same donor who had previously given stem cells underwent non-mobilized apheresis of peripheral blood followed by CD3 depletion and CD56 positive selection. The resulting product was incubated with a cytokine cocktail for 12-16 hours, washed, and infused into the lymphodepleted patient. Following NK cell infusion, low dose IL-2 was administered subcutaneously every other day for 7 doses. The clinical outcomes of patients treated with CIML NK cells are shown. All patients had measurable disease post haplo-HCT and prior to receiving CIML NK cells. Complete response and morphologic leukemia-free state are defined per the ELN 2017 criteria for AML, and marrow CR is defined as per the IWG criteria for MDS. **B** Next-generation sequencing was used to track individual mutations before and after treatment with the donor-derived CIML NK cells. Patients 2 and 3 exhibited marrow CR (for MDS) and CR without minimal residual disease (for AML) at the day+28 assessment, respectively, while Patient 1 exhibited a morphologic leukemia-free state with persistently detectable FLT3-ITD. Patient 5 had progressive disease as per ELN 2017 criteria for AML. **C** Disease response of the BPDCN patient who had no bone marrow involvement of disease at the time of CIML NK cell therapy. Shown is the PET/CT-imaging of the two identified lesions in the scalp with the corresponding visual evaluation of these lesions, and the active axillary lymph node.

All patients were evaluable at the day +28 response assessments following infusion of CIML NK cells, and 4 of 5 patients attained a response to therapy. Three of 5 patients met ELN 2017 criteria for CR in AML or IWG criteria for marrow CR in MDS [25, 26], including the patient with MDS and multiple pathogenic mutations who had no detectable residual disease by next-generation sequencing (Patient 2), the patient with AML and pathogenic TP53 mutation who had no detectable residual disease by next-generation sequencing (Patient 3) (**Figure 1B**), and the patient with BPDCN (Patient 4) (**Figure 1C**). Patient 4 had entirely extramedullary disease relapse prior to receiving CIML NK cell therapy, and his response was evaluated with PET/CT imaging. Patient 1 attained a morphologic leukemia-free state as she was able to clear some of the pathogenic mutations present at the time of post haplo-HCT relapse, but had a persistent FLT3-ITD mutation detectable at the day+28 assessment. Patient 5 had disease progression at the day+28 response assessment.

Two of the patients who attained CR following receipt of CIML NK cells had no detectable disease for 4 months (Patient 3) and 6 months (Patient 4) following infusion. Patient 2 died from septic shock due to fungal pneumonia 8 weeks after infusion of CIML NK cells, but had no evidence of disease relapse. While Patient 4 had entirely extramedullary disease at the time of CIML NK cell therapy, relapse of disease occurred in the bone marrow.

### Adoptive transfer of CIML NK cells is associated with rapid and prolonged expansion in the peripheral blood NK cell compartment without changes in Treg numbers

All CIML NK products consisted predominantly of subpopulations of CD3^-^CD56^+^ cells (**Figure 2A**). Following CIML NK cell infusion, NK cells dramatically expanded to a median peak of 10-fold (range 10-to 50-fold; **Figure 2B**). The increased NK cell numbers persisted for up to several months beyond the last dose of IL-2 on day +12. T-cells constituted only a minority of PBMCs in the peripheral blood compartment once the NK cells expanded, and CD3^+^CD8^+^ T-cells were the major subset of CD3^+^ T-cells during the first few weeks after CIML NK infusion (**Figure S1**). Exogenous administration of IL-2 in these patients did not significantly increase the absolute numbers of Tregs (**Figure 2C**). We correlated NK cell expansion with CMV reactivation and found that patients 1, 2, and 3 reactivated CMV following NK cell infusion, although with low viral loads and none needing CMV-directed treatments (**Figure S2**). Patients 4 & 5 did not have reactivation of CMV following CIML NK infusion.

**Figure 2.**
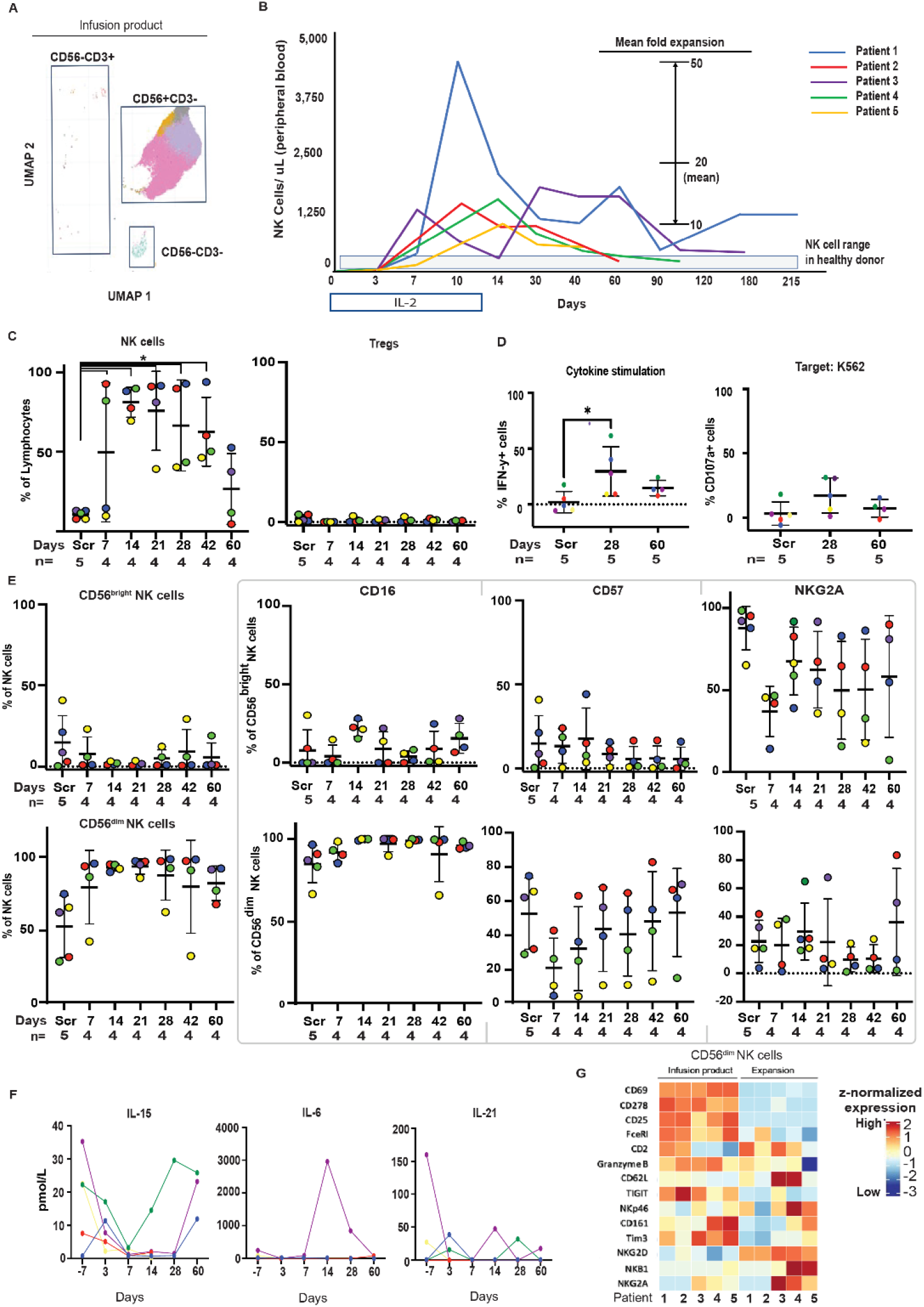
Rapid expansion of NK cells following infusion of CIML NK product into patients. **A** The infusion product in all patients is enriched with CD56^+^CD3^-^ NK cell populations. Shown is the UMAP of immune cell clusters derived from the infusion products of all 5 patients treated with CIML NK cell therapy. Individual subpopulations of CD56+CD3-cells are indicated with distinct colors. **B** Evaluation of peripheral blood mononuclear cells demonstrate a significant expansion of the NK cell compartment in all treated patients that persisted for several months after infusion. IL-2 was administered every other days for 7 doses starting Day 0. **C** Flow cytometry-based evaluation of NK cell numbers in the peripheral blood as a proportion of lymphocytes in all treated patients shows the dominance of the NK cell compartment until about Day+60 post CIML NK cell infusion. Also shown is the Treg subpopulation of total lymphocytes, characterized by CD3^+^CD4^+^CD25^+^CD127^-^ phenotype, demonstrating minimal expansion of these cells with exogenous administration of low dose IL-2. The number of samples at each time point, n=5, for screening, days 7, 14, 21, 28, while n=4 for days 42 and 60. This is because Patient 5 had disease progression by day +28 and came off study. The value for each patient is shown with a colored dot, and the coloring scheme corresponds to that in panel B. **D** Functional characterization of the expanded NK cell compartment using both cytokine stimulation and K562 target cells. The expression of CD107a and IFNγ was evaluated at the indicated time points shown, with the screening time point referring to endogenous patient NK cells prior to infusion, and the other time points being 28 days and 60 days after infusion of donor CIML NK. The y-axis refers to percent expression of the indicated marker relative to the corresponding unstimulated control. The color scheme for each patient data point corresponds to the same color scheme as in panel A. The dashed line represents the same functional assays applied to healthy donor control PBMCs. **E** Flow cytometry-based evaluation of key markers distinguishing the CD56^dim^ NK cell population from the CD56^bright^ NK cell population. Each of the colored dots represents a patient. The number of patients, n, at each indicated time point is indicated below the time point. **F** Measurement of plasma cytokines following CIML NK cell infusion. The x-axis of each plot refers to days relative to CIML NK cell infusion (Day 0). **G** Distribution of CD56^dim^ NK cell markers in all treated patients at Day +28 after CIML NK infusion compared to the CIML NK infusion products using mass cytometry. Patients are shown on the x-axis. Each row corresponds to the indicated markers on the CD56^dim^ NK cell populations. Relative expression is indicated, with red representing higher expression and blue lower expression. *Statistically significant comparisons using the Wilcoxon rank-sum test with p<0.05 are indicated.

Flow-based functional evaluation of infused NK cells at the screening, expansion (21-28 days) and post-expansion (60 days) time points was performed by cytokine stimulation or co-culture with K562 cells. At the expansion time point after CIML NK cell infusion, IFNg expression was elevated in response to cytokine stimulation (Wilcoxon signed rank p < 0.05; **Figure 2D**). While CD107α was upregulated after co-culture with K562 at the expansion time point compared to the screening time point in patients 1, 2, and 4, it was not in patients 3 & 5.

During their expansion, NK cells were predominantly CD56^dim^CD16^hi^ and displayed the classical NK cell maturity markers, including KIR and CD57 (**Figure 2E**). A subpopulation of CD3^-^CD56^+^ cells exhibiting the CD56^bright^ NK cell phenotype by flow cytometry was also noted in patients 4 & 5, particularly after day+28, although it remained relatively small. We evaluated the concentration of endogenous cytokines that have an association with NK cell proliferation, and noted that concentrations of IL-15 or IL-21 were not elevated at the time of exogenous IL-2 administration, although IL-15 did appear to increase in patient 4 after day+7 (**Figure 2F**). We noted that patient 3, the only patient who developed CRS after NK cell infusion, had a corresponding increase in IL-6 supporting the diagnosis.

We conducted a differential expression analysis of NK cell markers, detected by mass cytometry, comparing the CIML NK cells in the infused products to the expanded NK cells at day +28. Downregulated markers at expansion *in vivo* included CD69 and CD25, the latter absent by day +7 after infusion of CIML NK cells (**Figure S3**). Immunomodulatory markers such as Tim-3 and TIGIT are downregulated with expansion, while activating receptors such as NKp46 and NKG2D were upregulated. There is some heterogeneity with respect to NKG2A, whereby patients 3, 4, and 5 upregulate this receptor with expansion while patients 1 & 2 did not. The expression of Granzyme B, CD2, and CD62L appeared to be relatively lower on the CD56^dim^ NK cells at the time of expansion in patient 5, in contrast to the other patients (**Figure 2G**).

### Phenotypically defined subsets of infused CIML NK cells expand and persist long-term

We investigated the specific phenotypic imprint of the CIML NK cell preparation by comparing the CIML NK cell infusion product with the original resting donor-derived repertoire from screening samples. CIML NK cells exhibited a cytokine-stimulated signature, including a higher expression of CD25 and CD69, and a higher expression of CD161 that is thought to mark cytokine-responsive NK cells when compared to the resting donor NK cell repertoire [27]. Expression levels of NKG2D and TRAIL were lower on the infusion products from all patients compared to the screening time point. Similarly, Ki67 was also expressed at a lower level on the infusion product populations compared to the screening time point, suggesting that only a subset of NK cells in the infusion product actively proliferate after cytokine stimulation before receiving the IL-2 mediated survival signal. The immunomodulatory markers Tim-3 and TIGIT were also more highly expressed on the infusion product than at screening (**Figure S4**).

To gain a deeper characterization of NK cell subpopulations that expanded following infusion, CYTOF-defined clusters in the infusion products were generated with the following set of markers for each patient: CD56, CD3, NKG2A, IL-7Ra, KIR2DL1, NKG2C, NKp46, NKp30, Ki67. The resulting clusters represented a number of identifiable NK cell populations, including adaptive NK cells (NKG2C^+^KIR^+^), CD56^bright^ NK cells (CD56^high^NKG2A^high^KIR2DL1^-^) and CD56^dim^NKG2A^low^ NK cells (**Figure S5**). The infusion products in Patients 1 & 2 had a large subpopulation expressing both NKG2C and KIR, consistent with adaptive NK cells. Patients 3 & 4, on the other hand, had a subpopulation expressing a higher level of CD56 and NKG2A, and a relatively low amount of KIR and NKG2C, consistent with CD56^bright^ NK cells that were largely absent in the other patients.

Phenotypic tracking over time of the NK cell subpopulations present in the infusion product showed a persistence of some of the subpopulations up to day+28 after CIML NK infusion and beyond day +60. Patient 1 & 3 exhibited the longest persistence of NK cell subpopulations phenotypically matching those that were infused (**Figure 3A**). We applied single-cell RNA sequencing (scRNAseq) to further characterize the NK cells in these two patients in order to explore what transcriptionally defined subpopulations may have accounted for this long-term persistence (**Figure 3B**). Patient 1 had three identifiable NK cell subpopulations, two of which were consistent with adaptive NK cells based on the transcriptional profile (**Figure 3C**). Patient 3 had a transcriptionally defined adaptive NK cell cluster, but in contrast to Patient 1, also had a CD56^bright^ NK cell cluster whose presence was consistent with flow cytometry immunophenotypic data (**Figure 3D**). The remaining NK cell clusters in the two patients could not be definitively ascribed a single phenotype, and likely represented a mix of NK cell populations.

**Figure 3.**
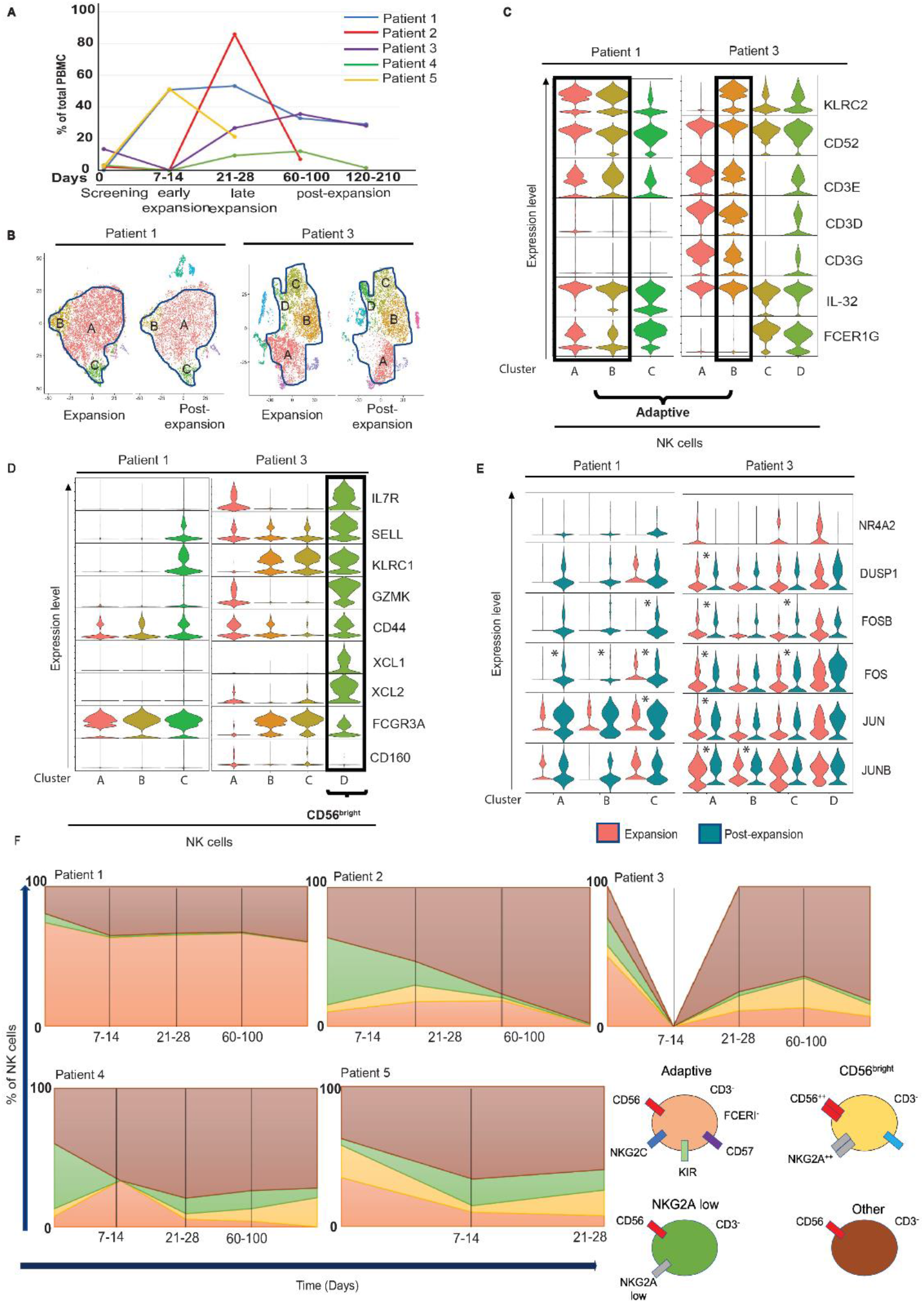
Tracking and evaluation of subpopulations of NK cells present in the infusion product over time. **A**. Median proportion of the CD56^+^CD3^-^ clusters that were present in the infusion product out of total PBMCs cells at the indicated time points. Days after infusion of the CIML NK product are indicated above the time point labels. **B** NK cell clusters defined with single-cell RNA sequencing at Days 21-28 (expansion) and days 60-210 (post-expansion) time points corresponding to Patients 1 & 3. At the post-expansion time points in both these patients, AML had relapsed. NK cell clusters are demarcated and labelled. **C** Identification of defined NK cell subpopulations based on transcriptional profiling reveals the expansion of predominantly adaptive NK cells (clusters A & B) based on the expression of NKG2C and other indicated markers in Patient 1. **D** Patient 3 is characterized by the expansion of a CD56^bright^ cluster of NK cells at the post-expansion time point, a cluster that is not identifiable in Patient 1. **E** Transcriptional profile of NK cell activation, with the vertical axis of the violin plots representing the expression level of the indicated transcript while the horizontal axis reflects the number of cells that express the transcript at the indicated level. The adaptive NK cell clusters in Patient 1 were not transcriptionally active at expansion (red). There were more cells at the post-expansion time point (green) that expressed markers of NK cell activation compared to the expansion time point. Differential expression of the indicated markers that meets the threshold for statistical significance with p< 0.05 is shown with an asterisk. The other NK cell populations in Patient 1 (cluster D) also exhibited a more transcriptionally active profile at the post-expansion compared to expansion time point. The CD56^bright^ cluster (cluster D) in Patient 3 exhibited a significant number of cells expressing transcriptional markers of activation at the expansion and post-expansion time points. **F** Tracking of NK cell subpopulations over time as a proportion of total NK cells in each of the clinical trial patients treated with CIML NK cell therapy. Key subpopulations being tracked are indicated in the legend, and include adaptive NK cells (CD56^+^CD3^-^CD57^+^KIR^+^NKG2C^+^FceRI^-^), CD56^bright^ NK cells (CD56^hi^CD3^-^NKG2A^+^IL7R^+^), a subpopulation of NKG2A low expressing cells (CD56^+^CD3^-^NKG2A^lo^), and all remaining NK cells. Time 0 of tracking corresponds to the screening time point of the trial before any CIML NK cells were infused.

While the adaptive NK cell clusters (clusters A & B) in patient 1 did not significantly upregulate genes associated with NK cell activation at the expansion time point (Day +28) and upregulated only a subset of these genes at the post-expansion time point (Day +60), the CD56^bright^ cluster (cluster D) in patient 3 exhibited a substantial number of cells expressing transcriptional markers of activation at both expansion and post-expansion points (**Figure 3E**). We further evaluated the transcriptionally-defined persistent adaptive and CD56^bright^ NK cell populations using CyTOF in all the trial patients (**Figure 3F**). We immunophenotypically confirmed the persistence of adaptive and CD56^bright^ NK cell populations in patients 1 & 3, in agreement with single-cell transcriptome data. We also noted the persistence of a CD56^bright^ NK cell population in patient 4 that appeared to expand after Day +28.

### The NK cell compartment undergoes a phenotypic shift with expansion, but reverts back to the screening phenotype at the post-expansion time point

CD56^dim^ NK cells remain a substantial immune subset in the peripheral blood at the post-expansion time point (**Figure 4A**). The CD56^dim^ NK cells post-expansion phenotypically resembled the corresponding NK cells at screening, with only NKG2D being differentially expressed (**Figure 4B**). Similarly, there were no markers that were significantly differentially expressed among CD56^bright^ NK cells between the two time points. The difference between the time points is mainly reflected in the greater abundance of the CD56^dim^CD3^-^CD57^+^CD16^+^KIR^+^ NK cell population at the post-expansion time point compared to screening in all evaluable patients (**Figure 4C**). We sought to identify the subpopulation of NK cells that might sustain this greater abundance by identifying a cluster among the NK cells that was highly positive for Ki-67 (**Figure 4D**). We noted that the relative abundance of this subpopulation post-expansion was significantly increased compared to the screening time point in all evaluable patients, although the number of Ki-67^+^ NK cells was < 1% of PBMCs at both time points (**Figure 4E**).

**Figure 4.**
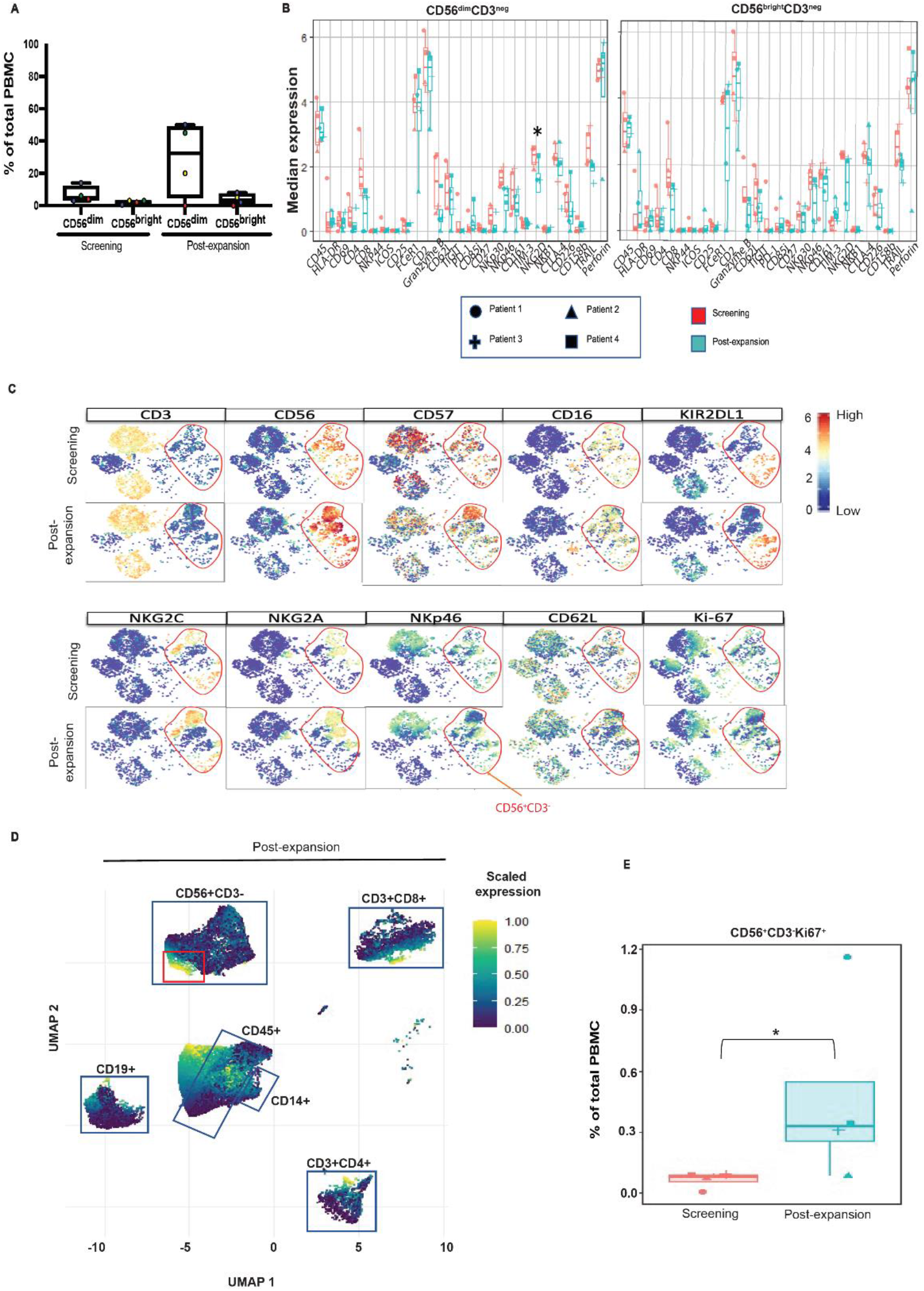
Phenotypic changes of the dominant NK cell subset in the peripheral blood over time. **A** Comparison of the screening and post-expansion time points relative to CIML NK infusion shows that at the latter time point there is a predominance of CD56^dim^ cells in the peripheral blood. At the screening time point, the median CD56^dim^ NK proportion of PBMCs was 3.3% (min 1.3%, max 15.2%), while the median CD56^bright^ NK proportion of PBMCs was 0.7% (min 0.1%, max 1%). At the post-expansion time point, the median CD56^dim^ proportion of PBMCs was 32.6% (min 0%, max 50.9%), while the median CD56^bright^ NK proportion of PBMCs was 2.5% (min 0 %, max 8.4%). **B** Differential expression of mass cytometry markers in both the CD56^dim^CD3^neg^ and CD56^bright^CD3^neg^ cell subpopulations identifies NKG2D as being the only marker whose downregulation on CD56^dim^ NK cells at post-expansion (green) relative to screening (red) among all patients meets statistical significance (Wilcoxon signed rank p < 0.05). The median marker expressions of the individual patients are shown. **C** Comparison of the screening and post-expansion time points reveals phenotypically similar populations of PBMCs, with a proportionally larger population of CD56^+^CD3^-^ cells at the latter time point. Shown is a representative tSNE plot from Patient 3. Markers indicated are associated with the memory-like phenotype of NK cells. NK cells are in the red circled area. **D** Clustering of PBMCs to identify the Ki67 subpopulation (red square) of CD56^+^CD3^-^ NK cells. Other clusters of immune cells are indicated. **E** The quantity of CD56^+^CD3^-^Ki67^+^ cells as a proportion of total PBMCs at both screening and post-expansion time points. Individual patient values are shown superimposed on the boxplots. The median percentage of CD56+CD3-Ki67+ cells at screening and post-expansion time points was 0.075% (min 0%, max 0.09%) and 0.32% (min 0.07%, max 1.15%), respectively. * corresponds to differential abundance of the indicated PBMC subpopulation between time points with p < 0.05 using Wilcoxon rank sum.

### NK cells traffic to sites of disease after CIML-NK cell infusion

We evaluated whether expansion of the NK cell populations after CIML NK cell infusion is associated with the presence of NK cells at sites of disease. Multiparameter immunofluorescence imaging (MIFI) allowed us to stain for multiple markers simultaneously in the bone marrow and tissue biopsy samples before and after CIML NK cell infusion. In Patient 4, CIML NK cell infusion was associated with an NK cell infiltrate at the tumor site 7 days post infusion. This infiltrate was juxtaposed next to CD123^+^ blasts, the burden of which was substantially reduced following infusion (**Figure 5A**). The reduction in blast burden was also associated with an influx of CD3^+^ cells into the tumor site. Further characterization revealed that >50% of these CD3^+^ cells at day +7 post infusion were also CD8^+^ (**Figure 5B**).

**Figure 5.**
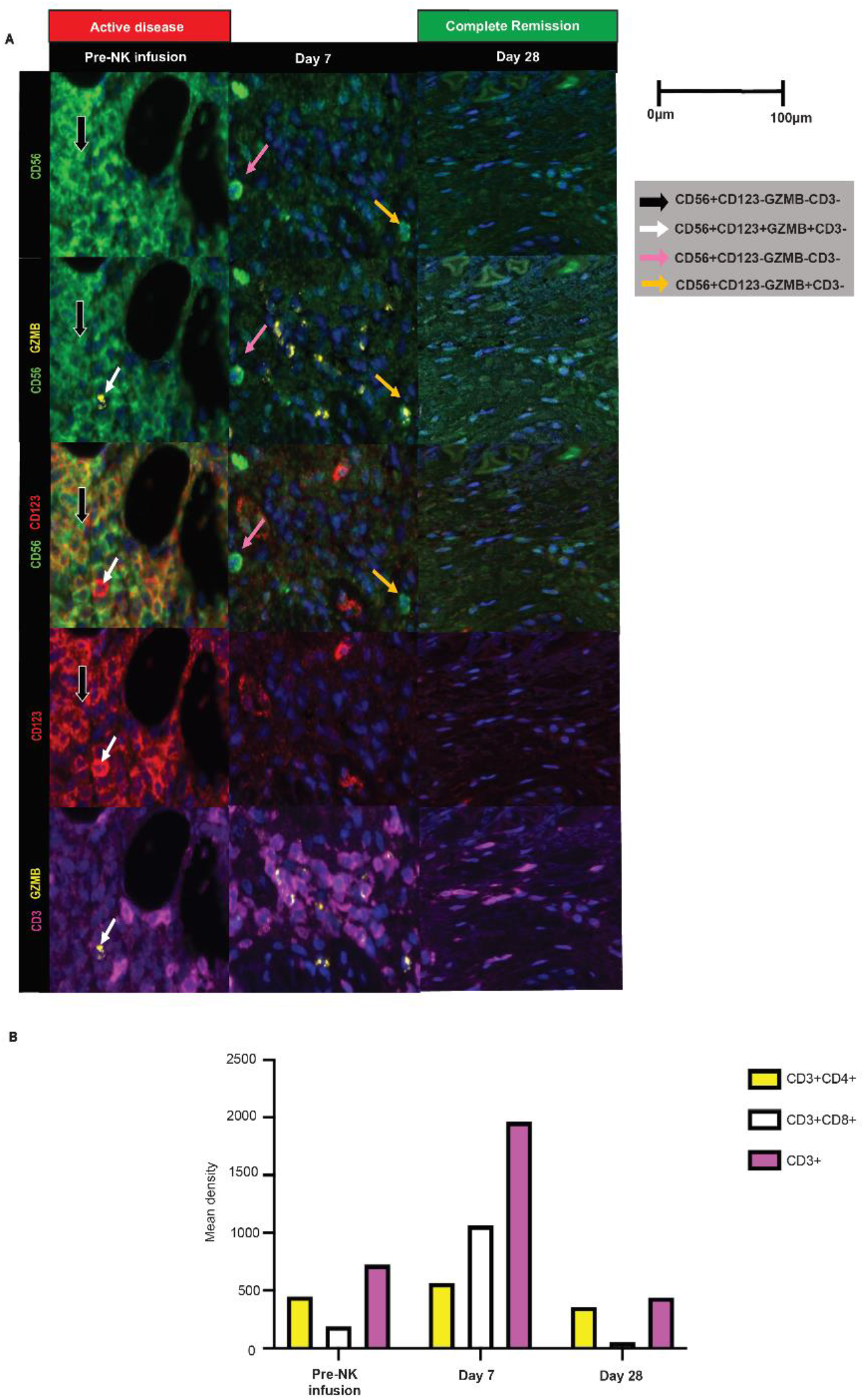
Detection of immune effector cells at the disease site. **A** Multiparameter immune fluorescence imaging of the extramedullary disease site in patient 4 demonstrates that juxtaposition of NK cells with disease cells is associated with significant reduction of disease burden. Longitudinal analysis of the extramedullary lesion prior to CIML NK cell infusion, 7 days post-infusion, and 28 days post infusion demonstrates that prior to infusion, there was a paucity of CD3^-^CD56^+^GranzymeB^+^ cells and CD3^+^ cells in the context of a dominance of CD56^+^CD123^+^ blasts. The black arrow highlights the presence of CD56^+^CD3^-^GranzymeB^-^ cells that are CD123^-^ and likely represent NK cells at the site of disease before CIML NK infusion. The white arrow indicates a blast cell that also expressed Granzyme B. Seven days after NK cell infusion, there was a substantial reduction in CD123^+^ blasts in the context of the presence of NK cells and an infiltrate of CD3^+^ cells. The pink and yellow arrows identify CD3^-^CD56^+^Granzyme B^-^ and CD3^-^ CD56^+^Granzyme B^+^ NK cells present at the site of disease, respectively. Most of the detected Granzyme B was associated with infiltrating CD3+ cells. At Day 28 after infusion, there remained complete absence of CD123^+^ disease. While there were persistent CD3^+^ cells, there were very few residual CD56^+^CD3^-^ cells remaining. **B** Measurement of the abundance of CD3^+^ infiltrate in the tissue based on mean fluorescence density. CD3^+^CD8^+^ cells exhibited the greater increase 7 days post NK cell infusion, eventually becoming undetectable 28 days after infusion.

In all other patients who had bone marrow involvement of their disease, MIFI demonstrated the presence of CD3^-^CD56^+^ cells in the marrow at days +21-28 post infusion (**Figure S6**). Patients 1 & 2 exhibited a predominantly empty bone marrow at the NK cell expansion time point, while patient 3 had a substantial amount of other immune cells, including CD3^+^ and CD8^+^ cells present concurrently with NK cells. Patient 5 had very few NK cells present in the bone marrow following expansion. Availability of screening, expansion, and post-expansion longitudinal bone marrow biopsy samples in patient 3 provided the opportunity to examine the changes that occur over time after infusion of CIML NK cells. At the post-expansion time point, there was a relative paucity of CD56^+^CD3^-^ cells. At the time of overt relapse whereby the peripheral blood contained circulating blasts, 1 month after administration of standard-of-care T-cell DLI and before any additional chemotherapy was administered (7 months post CIML NK infusion), there was a relative paucity of both CD3^+^CD56^-^ and CD3^+^CD8^+^ cells in the bone marrow (**Figure S6**).

### AML relapse post CIML NK cell therapy is associated with low expression of ligands for CD2

We observed that Patient 1 and Patient 3 both exhibited long-term persistence of NK cell populations that were present in their infusion products (**Figure 3A**), and at the time of AML relapse had a coexistence of leukemia blasts with the NK cells in the peripheral blood (**Figure 6A**). We conducted a differential expression analysis comparing NK cell clusters between expansion and relapse time points in both patients 1 & 3 to determine whether there was a change associated with relapse. All the NK clusters in Patient 1 had a significant reduction in the expression of *CXCR4* with the development of relapse (**Figure 6B**). Patient 3 on the other hand had an upregulation of genes associated with the CD56^bright^ phenotype of NK cells at the time of relapse. Gene set enrichment analysis identified the FGFR1c pathway, thought to be involved in the differentiation of CD56^bright^ NK cells to a CD56^dim^ phenotype, as being upregulated in the adaptive NK cell clusters of Patient 1 at the time of AML relapse relative to the time of NK cell expansion, while being downregulated in the CD56^bright^ cluster of Patient 3 when comparing the same two time points (**Figure 6C**).

**Figure 6.**
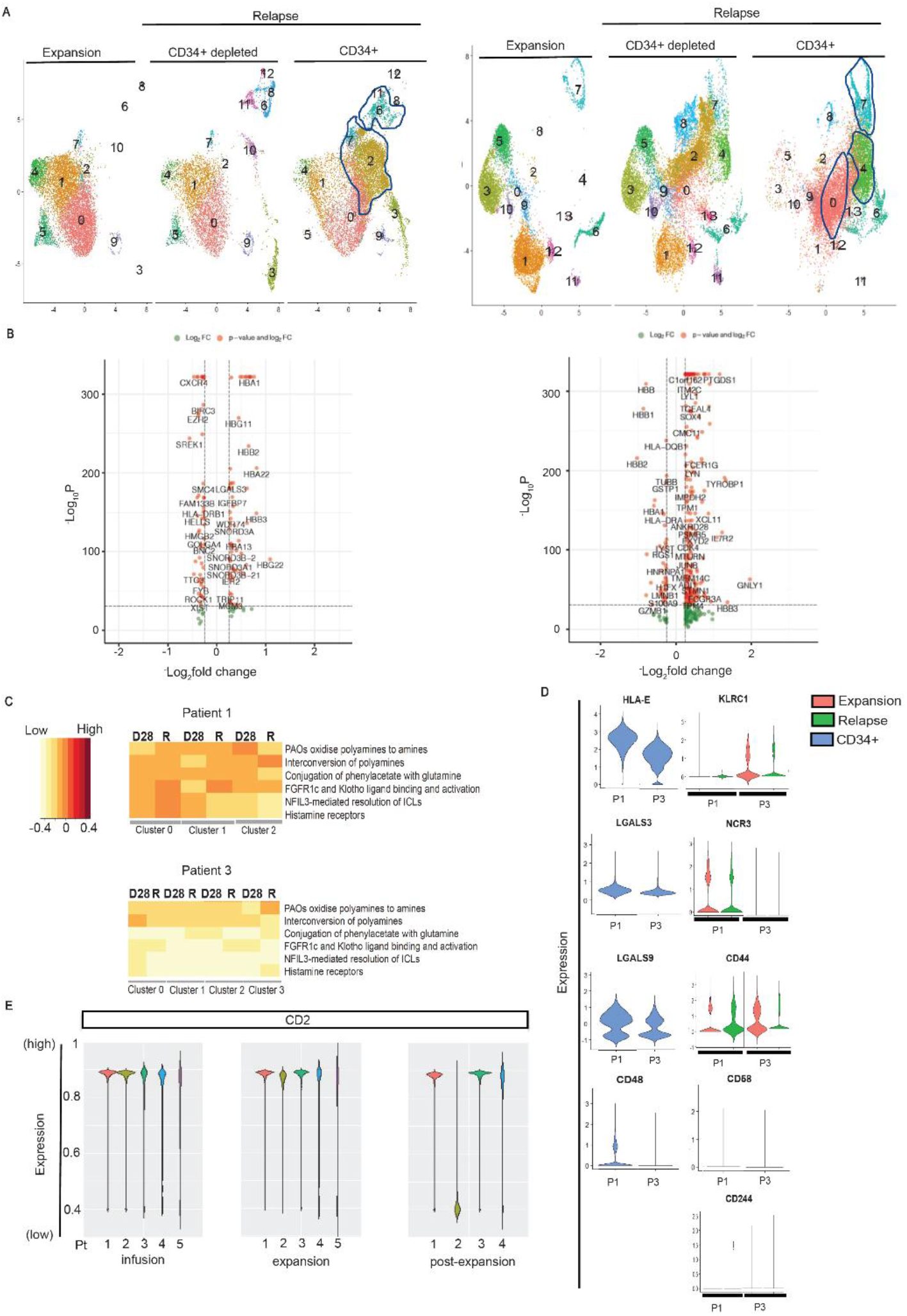
Single-cell RNA sequencing analysis of CD34^+^ enriched subset of cells at the time of relapse. **A** Identification of clusters of CD34^+^ cells at the time of relapse after CIML NK cell therapy. Expansion refers to day+28 after CIML NK infusion. The CD34+ fraction clusters that are enriched for acute leukemia blasts are demarcated in each patient. Clusters at the expansion time point are enriched for NK cells. **B** Volcano plots for Patient 1 (left) and Patient 3 (right) comparing gene expression in NK cell containing scRNAseq clusters at expansion to the corresponding clusters at relapse. Genes on the left of the violin plot are downregulated at relapse compared to expansion, while those on the right are upregulated. **C** Gene-set enrichment analysis corresponding to NK cell clusters derived from scRNAseq data in Patient 1 (top plot) and Patient 3 (bottom plot). Within every cluster, every pair of columns corresponds to expansion (D28) and relapse (R) time points, respectively. The intensity of expression of indicated pathways is represented by the color of the legend on the left. Gene sets that are differentially expressed between the two time points are shown. **D** Transcriptional profile of select NK cell receptors at the expansion (Exp) and relapse (R) time points. Red violin plots correspond to NK cell clusters at expansion, and green violin plots represent expression of markers at the corresponding NK cell clusters at relapse. The expression of activating and inhibitory ligands for the NK receptors on AML blasts at relapse is shown in Patient 1 (P1) and Patient 3 (P3) in the blue violin plots. CD48 and CD58 are NK activating ligands while LGALS9, LGALS3, and HLA-E are inhibitory ligands. CD244 is a receptor for CD48, CD44 is a receptor for LGALS9, NCR3 (NKp30) is a receptor for LGALS3, and KLRC1 is a receptor for HLA-E. The ligand for CD58, CD2, was not well transcribed in the scRNAseq data, but its expression is shown with mass cytometry in the next panel. The expression of other known NK ligands and receptors is shown in the supplementary appendix. **E** CD2 is expressed on NK cell containing clusters in patients at the indicated time points and persists after infusion. Expression is measured with mass cytometry applied to patient PBMCs at the following time points: screening (pre-CIML NK therapy), infusion (CIML NK product on day 0), early expansion (day +7-14 after CIML NK infusion), late expansion (day +21-28 after CIML NK infusion), and post-expansion (after day+60 post CIML NK infusion). Median scaled CD2 expression (y-axis) on NK cell clusters is indicated with box plots whose colors correspond to the patients as shown on the right.

We evaluated whether post-CIML NK relapse of AML was associated with the expression of ligands for both activating and inhibitory receptors present on the persistent NK cells in the peripheral blood. Using scRNAseq, we identified CD34^+^ containing clusters enriched for AML blasts, confirming that these did not express typical NK cells markers (**Figure S7**). Assessing the expression of NK ligands on the CD34^+^ clusters revealed that HLA-E, an inhibitory ligand that binds *NKG2A* and KLRD1 (CD94), was expressed highly on the CD34^+^ clusters in Patient 1 but its expression was lower on the CD34^+^ clusters in Patient 3 (**Figure 6D**). While the NK cells expressed CD2 stably through expansion and post-expansion (**Figure 6E**), the leukemia-containing clusters were found to express low levels of RNA for CD48 and CD58, both of which are activating ligands that bind CD2 (**Figure 6D**). Galectin-9 (LGALS9), a known inhibitory ligand for NK cells, is expressed predominantly on the CD34^+^ clusters in both patients. Tim-3 (*HAVCR2*), which is one of the receptors for Galectin-9, is not transcriptionally expressed on the CD34^-^ clusters at either the expansion or relapse time point, but another receptor, CD44, is expressed on the CD34^-^ clusters at relapse. The expression of most other NK activating and inhibitory ligands was either not present or not assessable with 3’ single-cell RNA sequencing (**Figure S8**).

## Discussion

We demonstrate for the first time that infusion of donor-derived CIML NK cells into an immune compatible environment for the management of post-haplo HCT relapse of myeloid disease is safe and results in rapid expansion and long-term persistence of NK cells. Infusion of CIML NK cells was associated with a reduction in measurable disease burden in 4 of 5 treated patients, including in a patient with BPDCN, with response to therapy sustained for several months. Furthermore, CIML NK cells were well-tolerated, with the main adverse effect being the development of one or more cytopenia in most patients. Importantly, infusion of these donor lymphocytes was not associated with severe CRS, neurotoxicity or GVHD.

The lifespan of conventional NK cells is thought to be in the range of 12-14 days, but adoptive transfer of CIML NK cells in a syngeneic murine model was associated with a significant prolongation of their expected lifespan [22]. In a previous clinical trial using CIML NK cells to treat relapsed/refractory AML in a non-transplant HLA mismatched setting, the donor-derived NK cells could not be detected beyond day +28 after infusion [20]. In the current clinical trial, expansion of donor CIML NK cells and subsequent persistence of these cells for several months was noted. We hypothesize that this was partly related to the immune-compatible post-transplant milieu since all treated patients had high T-cell chimerism at the time of CIML NK infusion. While upregulation of CD25 (IL2Ra) on the CIML NK cells in the infusion product could have supported the expansion and activation of the adoptively transferred cells in the presence of low dose IL-2, the prompt downregulation of CD25 seven days post-infusion suggests that other mechanisms must be involved in supporting long-term NK cell persistence.

Ki67 expression was relatively reduced in the infusion product, and expression was only increased in a small subset of NK cells at later time points. This may suggest that only a subpopulation of infused NK cells was responsible for expansion and long-term persistence. The expansion and persistence of this NK cell subpopulation in most patients was not associated with increased secretion of endogenous cytokines such as IL-15 and IL-21. CMV reactivation likely played a role in the expansion and persistence of NK cell subsets in three of the patients because it temporarily coincided with a spike in NK cell numbers in the peripheral blood. Furthermore, transcriptional profiling of the NK cells in one of these patients demonstrated the expansion and persistence of an adaptive NK cell population, supporting CMV as a potential driver of NK cell expansion in this patient.

The expanded NK cell compartment resulting from the infusion of CIML NK cells was associated with presence of NK cells at sites of disease involvement and reduction of disease burden. This was most dramatic in Patient 4 with BPDCN whose disease burden visibly shrank and became undetectable on PET/CT. Multiparameter immunofluorescence demonstrated the presence of both an NK cell and CD8^+^ T cell infiltrate in the extramedullary mass in this patient, suggesting a possible interaction between these two cell populations in mediating disease reduction and preventing disease relapse. The infiltration of CD8^+^ T-cells to the site of disease was also seen in Patient 3. Both of these patients had a sizeable CD56^bright^ NK cell population expand and persist in addition to the dominant CD56^dim^ NK cells that expanded in all patients. CD56^bright^ NK cells are predominantly associated with cytokine secretion. A plausible hypothesis may be that while the cytotoxic CD56^dim^ NK cells mediate an initial wave of disease burden reduction, the CD56^bright^ NK cells concurrently secrete cytokines that may be responsible for recruiting cytotoxic CD8^+^ T-cells into the tumor microenvironment in order to enhance the anti-leukemia effect. Further work will be required to explore this possible interaction between the CD8^+^ T-cells that appear to infiltrate disease sites and infused CIML NK cell subsets.

While the use of CIML NK cells was safe and not associated with severe CRS, neurotoxicity, or GVHD, it is of note that all patients with bone marrow involvement of their myeloid disease exhibited pancytopenia. In two of these patients, a CD34-selected stem cell boost was required to promote peripheral blood cell recovery while in the others the peripheral blood counts recovered on their own. One patient died as a result of severe sepsis in the context of prolonged pancytopenia post CIML NK infusion. Prolonged pancytopenia post adoptive cell transfer was observed in prior CAR-T trials and may be related to the lymphodepleting chemotherapy [28, 29]. It is also possible that the inflammatory changes induced locally in the marrow microenvironment following infusion of CIML NK cells may contribute.

While no patient developed severe CRS, one patient exhibited clinical and laboratory features of grade 2 CRS requiring treatment with tocilizumab after which the syndrome promptly resolved. Interestingly, NK cell expansion was dramatically disrupted at the time of elevated IL-6 levels coincident with clinical CRS, and rebounded shortly after administration of tocilizumab. The effect of IL-6 on NK cell proliferation has not been well-described, although there is evidence for IL-6 being associated with reduced NK cell cytotoxicity [30]. It is unclear whether the development of CRS significantly altered the ratio of CD56^dim^ to CD56^bright^ NK cells or resulted in any long-term effects on NK cell function in this patient after Tocilizumab therapy. The effect of high IL-6 secretion on CIML NK cell expansion and function merits further study.

CRS was also reported in a recently published haploidentical NK cell therapy trial, although the higher reported rate in that trial could have been due to the use of subcutaneous IL-15 to expand the NK cells [31]. Consistent with this trial and other published NK cell trials, no patient developed any GVHD in spite of receiving a product that is mismatched to the recipient. The tolerability of the adoptively transferred NK cell therapy in this trial is similar to what was seen in other NK cell therapy trials, including those using IL-21 expanded NK cells [32, 33].

The deeper immunophenotypic evaluation of infused CIML NK cells using mass cytometry revealed inter-patient heterogeneity, likely reflecting the diversity of the NK cell populations inherent in the donor [34]. Using mass cytometry to track changes in the phenotype of infused NK cells at various time points after infusion, it was demonstrated that the CIML phenotype persists long-term, as an imprint superimposed on the original NK cell repertoire. However, by around day+60 post-infusion, the NK cell subpopulation distribution largely resembles that seen prior to infusion of CIML NK cells. The main difference refers to a larger size of the post-infusion NK compartment compared to the study screening time point.

Single cell RNA sequencing analysis of the NK cell compartment at the time of AML relapse post CIML NK cell therapy demonstrated the persistence of both CD56^bright^ and adaptive NK cell populations among others. Gene set enrichment analysis suggested that in the patient with a sizeable expansion and persistence of the CD56^bright^ NK cells, the NK cells exhibited downregulation of the FGFR1c pathway involved in NK cell maturation from CD56^bright^ to CD56^dim^, potentially explaining why the CD56^bright^ NK cells were able to persist [35]. The expression of HLA-E in the relapsed AML blasts in this patient with an expanded NKG2A-expressing CD56^bright^ NK population suggests a potential evasion mechanism, in agreement with previous analysis of residual leukemic cells following allogeneic NK cell therapy [36]. Since this patient also exhibited an infiltrate of CD8^+^ T-cells at the site of disease, possibly promoted by the cytokine secretion from the CD56^bright^ NK cells, evasion of CD8^+^ T-cells may have contributed significantly to the relapse mechanism.

Additional potential mechanisms of AML relapse post CIML NK therapy may be associated with the absence of ligands for CD2, a receptor that is expressed on the NK cell populations after infusion, through expansion and post-expansion, and whose interaction with CD16 is involved in mediating tumor target cytotoxicity [37]. In Patient 1 in particular, where most of the expanded and persistent NK cells exhibited a mature cytotoxic CD56^dim^CD16^+^ phenotype, the absence of expression of CD48 and CD58 on relapsed leukemia could suggest a resistance mechanism. Other mechanisms of AML relapse may involve the inhibitory ligand Galcetin-9, since its receptor CD44 [38] was expressed on the CD34^-^ peripheral blood mononuclear cells at the time of relapse. The expression of Galectin-9 may have played an additional role in the survival of residual leukemia clones early after NK cell infusion when its other ligand Tim-3 was more highly expressed in the infused CIML NK cell product.

While this study consistently demonstrated the rapid expansion and persistence of donor-derived CIML NK cells without any GVHD or severe CRS complications, it has important limitations. First, the number of treated patients is still relatively small. Furthermore, as the infused NK cells share HLA expression with the NK cells already present in the transplanted patient by virtue of coming from the same donor, it is not possible to use donor chimerism as a means to track infused NK cells. Although these limitations prevent making immediate generalizations of the findings, the deep high-throughput analyses of longitudinally collected samples do reveal a number of important hypothesis-generating observations with the potential for clinical translation in the near future. Additional work is necessary to elucidate the mechanism(s) of the prolonged alterations in the NK cell compartment after donor CIML NK cell infusion as well as their interaction with other immune effector cells as a means to treat post-transplant relapse of myeloid disease.

## Conclusion

In summary, we demonstrate that CIML NK cells infused into an immune-compatible milieu in the context of a haploidentical stem cell transplant are associated with marked NK expansion and long-term persistence. We show that CIML NK cell products are composed of predominantly cytotoxic CD56^dim^ NK cells but also exhibit donor-specific heterogeneity in both the presence of additional NK cell subsets as well as in the expression of NK cell markers associated with the memory-like phenotype. The cytokine-stimulated NK cell subpopulations present at the time of NK cell infusion undergo significant phenotypic changes associated with enhanced NK cell activity and the development of a memory-like phenotype by Day +28, but by about Day +60 post-infusion resemble phenotypically the NK cells present prior to infusion despite being present in significantly increased numbers. And finally, we speculate that relapse of AML after CIML NK cell therapy may occur in the context of evasion of CD2-mediated activation. Because of their capacity for rapid expansion and long-term persistence, CIML NK cells offer an attractive platform for further improvement of NK cell therapies by combining them with novel immunomodulatory agents and or by arming them with CAR gene constructs.

## Methods

### Study Design

Patients with relapsed myeloid disease, including acute myeloid leukemia (AML), myelodysplastic syndrome (MDS), myeloproliferative neoplasm (MPN), or blastic plasmacytoid dendritic cell neoplasm (BPDCN) following haplo-HCT were eligible to be treated on a single center phase I dose de-escalation clinical trial of CIML NK cell therapy (*NCT04024761*). The primary endpoints are safety and identification of the maximum tolerated dose (MTD) of CIML NK cells administered following stem cell transplantation. CIML NK cells were derived from the same donor who provided stem cells for the original transplant. Patients receiving therapy had relapsed disease that was confirmed by morphological evaluation, flow cytometry, next-generation sequencing, or PET/CT imaging, did not previously receive DLI, and had to have at least 20% donor T-cell chimerism. NK cells were purified from non-mobilized apheresis products by a two-step CD3^-^ depletion followed by CD56^+^ selection (CliniMACS^®^ device, Miltenyi Biotec), consistently achieving >90% purity for CD3^-^CD56^+^ NK cells. The purified NK cells were subsequently activated overnight (12-16 hours) with IL-12 (10ng/ml), IL-15 (100ng/ml), and IL-18 (50ng/ml) to generate the CIML NK cells under GMP conditions. The cells were washed and were subject to quality control assessment including a requirement for a T cell dose of < 3×10^5^ cells/kg in the product. All patients were infused at an NK cell dose of 5-10 × 10^6^ cells/kg after a course of lymphodepleting chemotherapy. Patients 1-4 were treated with a combination of fludarabine (25 mg/mg^2,^ days −6, −5, −4, −3, −2), and cyclophosphamide (60mg/kg, given on days −6 and −5), while the lymphodepleting chemotherapy doses for Patient 5 were adjusted to fludarabine (30mg/m^2^ IV over 1 hour on days −5, −4, −3) and cyclophosphamide 500mg/m^2^ IV over 1 hour (on days −5, −4) after a trial amendment. All subjects were treated with a low dose human recombinant IL-2 (1 × 10^6^ IU/m^2^) subcutaneously every other day for a total of 7 doses. Response evaluation included day +28 bone marrow biopsy and a PET / CT in patient with extramedullary disease (patient with BPDCN).

### Patient and healthy donor samples

Patients with relapsed AML, MDS, MDS/MPN, or BPDCN provided written informed consent prior to participation under an IRB-approved protocol at the Dana-Farber Cancer Institute. Correlative samples included peripheral blood collected at screening for the trial, on day −7 (before starting lymphodepletion), day 0, day +7, day +21-28, day +42, day +60, day +100, and relapse time points after infusion. Healthy donor PBMCs (peripheral blood mononuclear cells) were isolated by Ficoll centrifugation, and NK cells were purified using RosetteSep (STEMCELL Technologies). All PBMCs were cryopreserved according to a standard internal laboratory protocol at the time of collection from patients.

### Flow cytometric analysis

A custom NK cell panel of antibodies was used (**Table S2**). All cell staining for flow cytometry was performed as previously described [20], and data were acquired on a BD LSR Fortessa flow cytometer and analyzed using FlowJo (Tree Star) software.

### Mass cytometry (CyTOF)

Metal-tagged antihuman antibodies used for our mass cytometry panel are listed in **Table S3**. In-house conjugations of antibodies were performed with Maxpar labeling kit per manufacturer instructions (Fluidigm). All antibodies were used per the manufacturer’s recommendation.

Cryopreserved patient PBMC samples were thawed, counted using AO/PI and pelleted by centrifugation at 1800RPM for 3 minutes. Cells were then incubated in 103Rh viability stain for 15 minutes, washed in CyFACS and incubated with undiluted Human TruStain FcX for 10 min for Fc receptor blocking. Antibodies mastermix was applied to cell suspension for 30 minutes, washed and fixed/permeabilized with FoxP3 Fixation/Permeabilization Concentrate and Diluent, prepared following manufacturer’s guidelines (eBioscience). A mix of intracellular antibodies prepared with 1X Perm Wash was added to each sample and incubated for 30 minutes. Next, cells were washed with 1X Perm Wash and incubated overnight at 4°C in FoxP3 Fixation/Permeabilization Concentrate and Diluent, containing 191/193Ir DNA Intercalator. Prior to acquisition, samples were transferred to 5 mL round-bottom polystyrene tubes with cell strainer caps, washed and filtered with Cell Staining Buffer (CSB), Cell Acquisition Solution (CAS), and resuspended in CAS supplemented with EQ Four Element Calibration Beads (1:10).

All mass cytometry data was collected on a HeliosTM Mass Cytometer (Fluidigm). The instrument was tuned using CyTOF Tuning Solution according to the Helios User Guide (Fluidigm, p. 60-68). A brief overview of tuning steps includes Pre-XY Optimization, Mass Calibration, XY Optimization, DV Calibration, Dual Calibration, Gases/Current Calibration, and QC report. EQ Four Element Calibration Beads (1:10 in CAS) were used according to the manufacturer protocol before and during acquisition. The data was normalized using the FCS Processing tab of the Fluidigm CyTOF Software 7.0.8493.

Data analysis was manually performed using FlowJo 10.7.1. Initial data cleanup was based on the gating strategy previously described [39]. Cell events were gated to remove dead cells and debris through biaxial plots of Time vs. Event Length, Beads (for removal of the EQ Calibration Beads), and Gaussian-derived parameters (Residual, Width, Offset). The viability stain 103Rh, was used to gate out dead cells on both K562 and PBMC populations. All viable cells were backgated on both DNA parameters (191Ir and 193Ir) to ensure no doublets were included. Clustering of NK cell populations was performed both with R-Phenograph and FlowSOM methods in R [40, 41].

### Luminex cytokine assays

Patient plasma samples were thawed and prepared for soluble analyte assay according to previously published methods [42, 43]. Analytes were measured on a Luminex FLEXMAP3D (Luminex, Austin, TX) per the manufacturer’s protocol. Soluble Eotaxin, GM-CSF, GROα, IFNα, IFNγ, IL-1β, IL-1α, IL-1RA, IL-2, IL-4, IL-5, IL-6, IL-7, IL-8, IL-9, IL-10, IL-12 p70, IL-13, IL-15, IL-17A, IL-18, IL-21, IL-22, IL-23, IL-27, IL-31, IP-10, MCP-1, MIP-1α, MIP-1β, RANTES, SDF1α, TNFα and TNFβ were tested. Of all soluble markers measured, 33 were within detectable range and could be quantified by extrapolation of MFIs to the respective standard curve between lower limit of quantitation and upper limit of quantitation. Analyte concentration for each patient was calculated using standard curves. Fold changes were calculated as a ratio relative to the patient’s baseline (C0D1) [44-46].

### Functional and flow-based cytotoxicity assays

Frozen PBMCs were thawed and cultured overnight in RP10 medium (RPMI 1640 supplemented with 10% FBS, 1× penicillin/streptomycin, 2mM L-glutamine, and 7.5 mmol HEPES) with 1ng/mL IL-15. K562 cells were cultured in RP-10 medium and labelled with 5uM of CellTrace Violet (Thermo Fisher Scientific) in PBS for 20 min at 37°C. PBMCs and K562 were washed twice with RP-10 and co-cultured at the indicated E:T ratios. To measure NK cell cytotoxicity, PBMCs and K562 cells were co-cultured for 4h, then stained with PE-Annexin V and 7-AAD (BD Biosciences) for 15 min at RT. To measure intracellular IFNγ and degranulation, PBMCs and K562 cells were co-cultured for 1h, followed by the addition of 0.2uL BD GolgiPlug™ (BD Biosciences), 0.13uL BD GolgiStop™ (BD Biosciences) and 1uL of APC-CD107a (Biolegend). After an additional 5h of co-culture, cells were stained for intracellular IFNγ using BD Cytofix/Cytoperm (BD Biosciences). Cells were acquired using BD LSR Fortessa and analyzed using FlowJo (Tree Star).

### Single-cell RNA sequencing and analysis

Cryopreserved PBMC samples from patients were thawed and diluted with pre-warmed complete RPMI, and cells were centrifuged at 1000rpm for 10 min to remove dead cells. Cell pellets were resuspended in 25% Percoll solution in PBS and centrifuged at 1250rpm for 10min to further remove dead cells. Live cells were resuspended in 2%FBS PBS for single cell isolation and cDNA library construction. The relapse samples were CD34-depleted using a CD34^+^ positive selection kit (STEMCELL technologies), with the CD34^+^ cells resuspended as independent tumor samples.

Single-cell libraries were prepared using the 10x Chromium Next GEM Single Cell 3’ Kit (10XGenomics), according to the manufacturer’s instructions. The single-cell cDNA libraries were sequenced by NovaSeq S4 flowcell (Illumina). Raw sequences were demultiplexed, aligned, and filtered. Barcode counting and unique molecular identifier (UMI) counting was done with Cell Ranger software v3.1 (10XGenomics) to digitalize the expression of each gene for each cell. Analysis was performed using the Seurat 3.0 package [47]. Data from individual samples was analyzed separately prior to combining the data from multiple samples. The outlier cells with very low number of gene features (<500) or low total UMI (<1000), or high number of gene features (>5000) or high total UMI as doublets (>20000) were removed. Cells with high mitochondrial ratio (>15%) from each data set were also removed. Subsequently, samples were combined based on the identified anchors for the following integrated analysis. Principal component analysis (PCA) was done and the first 15 principal components (PCs) were used to perform tSNE clustering. Well-defined marker genes for each cluster were used to identify potential cell populations, such as T cells (CD3E, CD4, CD8A), B cells (CD19, CD20, SDC1), macrophages (CD14), and NK cells (NCAM1, FCγRIII, Granzyme B). For gene sets representing specific cellular functions or pathways, we performed functional enrichment analysis with the online tool Reactome GSA (https://www.biorxiv.org/content/early/2020/04/18/2020.04.16.044958) [48].

### Statistical Analyses

All flow cytometry data were tested for normal distribution (Shapiro-Wilks test), and if the data were not normally distributed, the appropriate non-parametric tests were used (GraphPad Prism v5.0). p-values corresponding to the differential expression using mass cytometry data were acquired using the *diffcyt* package in R [34]. Differential expression of single-cell RNA sequencing data was performed using the Wilcoxon rank-sum test as part of the Seurat package [32]. All statistical comparisons are indicated in the figure legends. All comparisons used a two-sided alpha of 0.05 for significance testing.

## Data Availability

De-identified correlative study data used for all analyses and to generate the figures is available upon request with the permission of the corresponding author.

## Study Approval

This study was reviewed and approved by the institutional review board of the Dana-Farber Cancer Institute, Boston, MA, USA (Clinicaltrials.gov identifier: *NCT04024761*). The study was performed in compliance with the provisions of the Declaration of Helsinki and Good Clinical

Practice guidelines. Written informed consent was obtained from participants before inclusion in the study.

## Author Contributions

RMS, SN, RJS, JR, and RR designed the clinical study protocol. RMS and RR designed the correlative studies. SN prepared CIML NK products under good manufacturing practice (GMP) guidelines and was responsible for quality control of the products. CR and MH processed and prepared patient samples for correlative studies. JV, AA, and MT performed the NK cell functional assays. BR and YA performed flow cytometry. GH performed single-cell RNA sequencing. JB performed mass cytometry. SR and NC performed multiparameter immunofluorescence on bone marrow biopsies. AAL provided BPDCN patient samples. CSC, JHA, VTH, JK, and MG recruited clinical trial participants. RCL and HTK reviewed the data from correlative studies and statistical analyses, respectively, and provided critical feedback. RMS and GB analyzed all the data and designed the figures. RMS, GB, and RR interpreted the data and wrote the first draft of the manuscript. KJM, CJW, JC, RJS, and JR reviewed the manuscript, and provided critical feedback on the results of the correlative studies. All authors revised the manuscript critically and approved the final version.

## Acknowledgements

We would like to thank M. Shipp for discussion and guidance, and D. Hearsey and C. Reynolds for technical assistance in preparing and processing clinical samples. We are grateful for the patient volunteers who gave their time and provided the samples for all correlative studies. We are also grateful to the bone marrow transplant and research coordinators at the Dana-Farber Cancer Institute. We acknowledge the use of the Dana-Farber Cancer Institute core for CyTOF mAb conjugation, and the Massachusetts Institute of Technology sequencing facility for single-cell RNA sequencing. We also acknowledge all the technicians at the Dana-Farber Cancer Institute cell processing facility for their work in generating clinical grade CIML NK cells for infusion.

