## Supplemental Tables and Figures for "Expansion, persistence and efficacy of donor memory-like NK cells for the treatment of post-transplant relapse"

**Supplementary Data**

**Table S1.** Clinical characteristics of trial patients prior to receiving CIML NK cell therapy

| Patient | Diagnosis | Treatment prior to CIML NK |
|---|---|---|
| Patient 1 | Secondary AML with FLT3-ITD | (07/18) 3+7 + Midostaurin<br>(10/18) RIC PBSC HaploHCT<br>(06/19) Relapse<br>(07/19) Gilteritinib<br>(09/19) added Decitabine for persistent disease, no response<br>(10/19) added Venetoclax for persistent disease, no response |
| Patient 2 | Secondary AML with TP53 mutation, multiple other pathogenic mutations | (08/18) 3+7<br>(12/18) RIC PBSC HaploHCT<br>(02/20) Relapse |
| Patient 3 | MDS with excess blasts II, multiple pathogenic mutations | (03/19) AZA x 2<br>(06/19) AZA + Venetoclax x 2<br>(11/19) RIC PBSC HaploHCT<br>(06/20) Relapse |
| Patient 4 | BPDCN | (07/19) Tagraxofusp<br>(09/19) RIC PBSC HaploHCT<br>(07/20) Relapse |
| Patient 5 | Secondary AML with multiple pathogenic mutations | (08/16) 3+7 + GMI-1271 clinical trial<br>(02/17) RIC DUCBT<br>(07/18) Relapse<br>(07/18) IMGN362 clinical trial<br>(09/18) Merestinib + LY2874455 clinical trial<br>(12/19) Decitabine + Venetoclax<br>(06/19) RIC PBSC HaploHCT<br>(12/20) Relapse |

**Table S2.** Flow cytometry antibody panels

**Tube 1 T CELLS**

| Antibody | Fluorochrome |
|---|---|
| CD25 | FITC |
| PD1 | PE |
| CD95 | PeCF594 |
| TCR a/b | PeCy7 |
| CD127 | APC |
| CD8 | AF700 |
| TCR g/d | APC Vio770 |
| TIM3 | BV421 |
| CD4 | BV510 |
| CD45RA | BV605 |
| CD6 | BV650 |
| CCR7 | BV711 |
| CD3 | BV786 |

**Tube 3 NK CELLS**

| Antibody | Fluorochrome |
|---|---|
| CD16 | FITC |
| PD1 | PE |
| CD95 | PeCF594 |
| NKG2D | APC |
| CD8 | AF700 |
| CD3 | BV786 |
| CD56 | BV605 |
| CD6 | BV650 |
| CD57 | PerCP Cy5.5 |
| NKG2A | BV421 |
| KIRs | PeCy7 |

**Table S3. Mass Cytometry Panel Antibodies for NK Cell Phenotyping Assessment.**

| **Antibodies** | Source | Cat. no. |
|---|---|---|
| **Anti-Human CD45-89Y** | Fluidigm | 3089003B |
| **Purified anti-human HLA-DR (Maxpar® Ready) Antibody*** | Biolegend | 307651 |
| **Anti-Human CD3-141Pr** | Fluidigm | 3141019B |
| **Anti-Human CD19-142Nd** | Fluidigm | 3142001B |
| **Anti-Human CD127[IL-7Ra]-143Nd** | Fluidigm | 3143012B |
| **Anti-Human CD69-144Nd** | Fluidigm | 3144018B |
| **Anti-Human CD4-145Nd** | Fluidigm | 3145001B |
| **Anti-Human CD8a-146Nd** | Fluidigm | 3146001B |
| **Anti-Human TNFα-146Nd** | Fluidigm | 3146010B |
| **Ultra-LEAFTM Purified anti-human CD336 (NKp44) Antibody*** | Biolegend | 325121 |
| **Anti-CD278/ICOS-148Nd** | Fluidigm | 3148019B |
| **Anti-Human CD25-149Sm** | Fluidigm | 3149010B |
| **Anti-Human FceRI-150Nd** | Fluidigm | 3150027B |
| **Anti-Human CD107a-151Eu** | Fluidigm | 3151002B |
| **Anti-Cleaved Caspase 3 (D3E9)-142Nd** | Fluidigm | 3142004A |
| **Granzyme B Antibody*** | Novus Bio | NBP1-50071 |
| **Anti-Human CD62L-153Eu** | Fluidigm | 3153004B |
| **Anti-Human TIGIT-154Sm** | Fluidigm | 3154016B |
| **Anti-Human CD279/PD-1-155Gd** | Fluidigm | 3155009B |
| **Anti-Human CD85j-156Gd** | Fluidigm | 3156020B |
| **Anti-Human CD27-158Gd** | Fluidigm | 3158010B |
| **Anti-Human CD33-158Gd** | Fluidigm | 3158001B |
| **Anti-Human CD337/NKp30-159Tb** | Fluidigm | 3159017B |
| **Anti-Human CD14-160Gd** | Fluidigm | 3160001B |
| **KIR2DL1/CD158a Antibody*** | Novus Bio | NBP2-11758 |
| **Anti-Human CD335/NKp46-162Dy** | Fluidigm | 3162021B |
| **Anti-Human CD56-163Dy** | Fluidigm | 3163007B |
| **Anti-Human CD161-164Dy** | Fluidigm | 3164009B |
| **Anti-Human IFNγ-165Ho** | Fluidigm | 3165002B |
| **Purified anti-human CD366 (Tim-3) (Maxpar® Ready) Antibody*** | Biolegend | 345019 |
| **Anti-Human CD314/NKG2D-166Er** | Fluidigm | 3166016B |
| **Anti-Human CD158e1/NKB1-167Er** | Fluidigm | 3167013B |
| **Anti-Human Ki-67-168Er** | Fluidigm | 3168007B |
| **Anti-Mouse CD8a-168Er** | Fluidigm | 3168003B |
| **Anti-Human CD159a/NKG2A-169Tm** | Fluidigm | 3169013B |
| **Anti-Human CD152-170Er** | Fluidigm | 3170005B |
| **Anti-Human CD226-171Yb** | Fluidigm | 3171013B |
| **FITC Antibody*** | Bio-Rad | 640001 |
| **Human NKG2C/CD159c Antibody*** | R&D | MAB1381 |
| **Anti-Human CD158b-173Yb** | Fluidigm | 3173010B |
| **TRAILR4/TNFRSF10D/DcR2 Antibody - Azide Free*** | Novus Bio | NBP1-45027 |
| **Anti-Human MIC A/B (6D4)-174Yb** | Fluidigm | 3174016B |
| **Anti-Human Perforin-175Lu** | Fluidigm | 3175004B |
| **Anti-Human CD57-176Yb** | Fluidigm | 3176019B |
| **Anti-Human CD16-209Bi** | Fluidigm | 3209002B |

***Denotes antibodies conjugated in-house.**

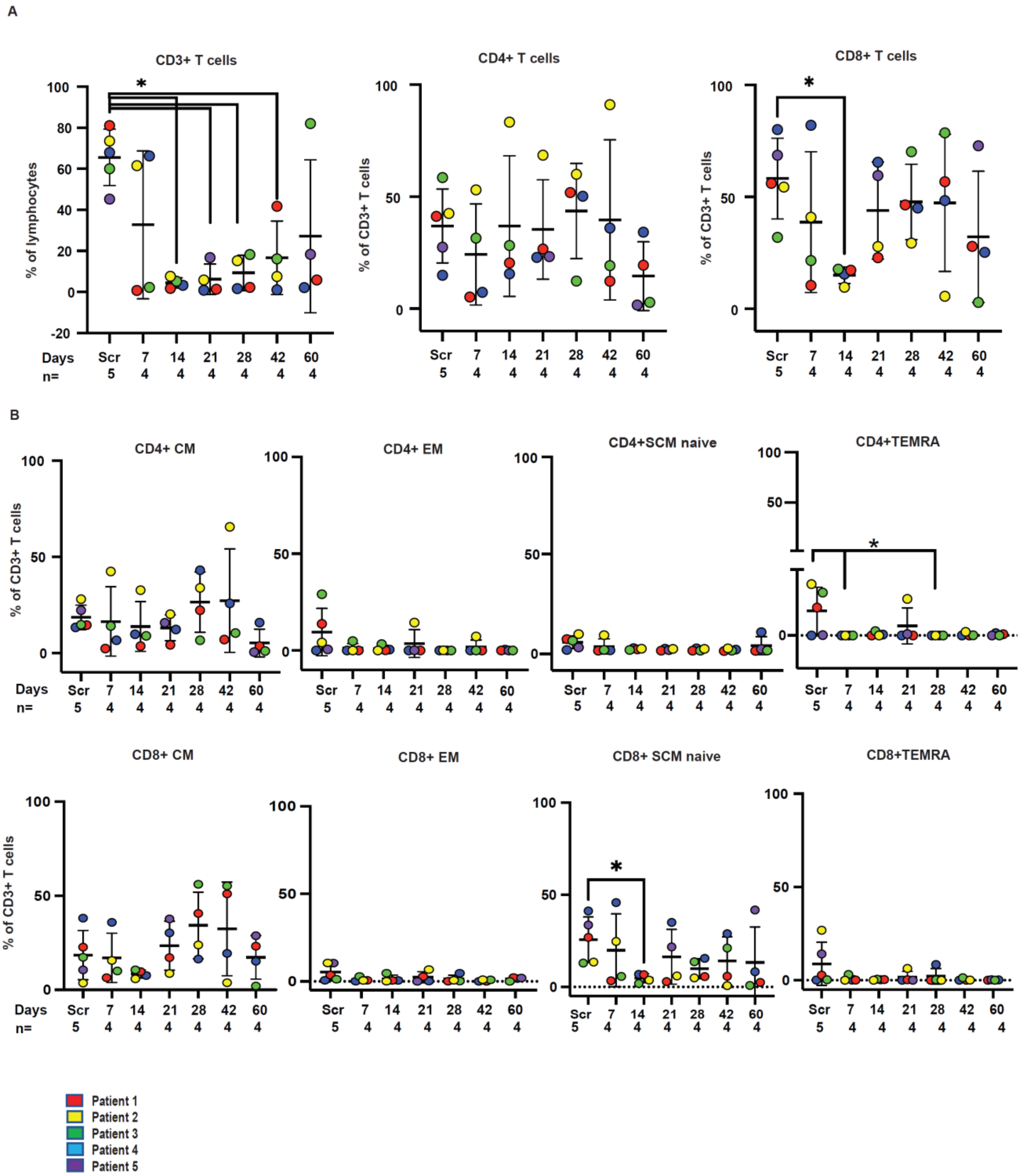


**Supplementary Figure S1.** Distribution of T-cell populations at various time points post-CIML NK cell infusion. The individual colors of all data points correspond to the patients labelled at the bottom. **A** Shown are CD3$^{+}$, CD4$^{+}$, and CD8$^{+}$ T-cells as a percentage of total lymphocytes at the indicated time points, including at screening prior to CIML NK cell infusion and at days +7, +14, +21, +28, +42, and +60 after infusion. **B** Further characterization of T-cell subsets, including CD4$^{+}$ and CD8$^{+}$ central memory (CM), effector memory (EM), memory stem cells (SCM), and effector memory T-cells expressing CD45RA (TEMRA).

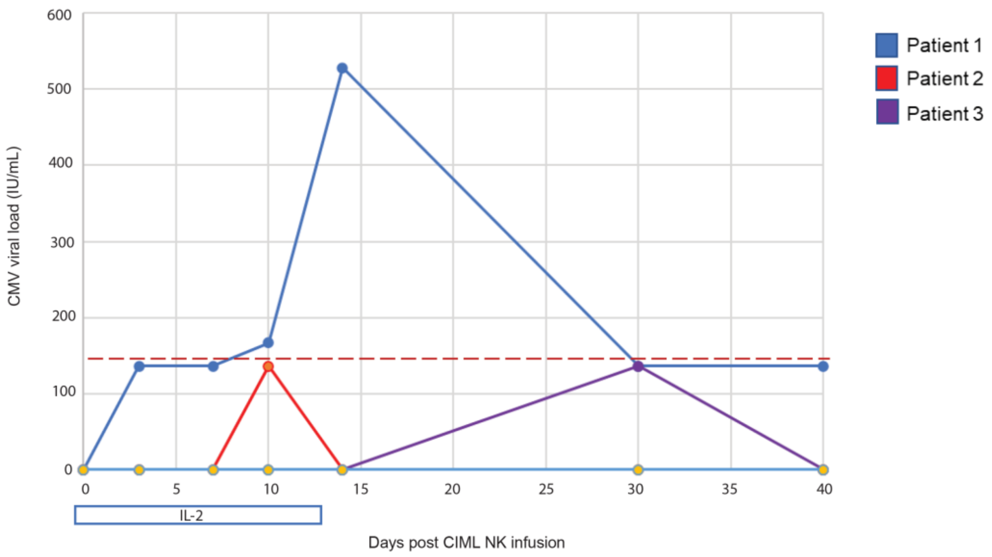


**Supplementary Figure S2.** CMV reactivation post CIML NK cell infusion in treated patients. Only patients 1, 2 & 3 are shown as CMV did not reactivate to any measurable degree in patients 4 & 5. Day 0 on the x-axis refers to the date of CIML NK infusion. The red dashed line indicates the lower limit of quantitation of the CMV viral load, although the virus is detectable.

A

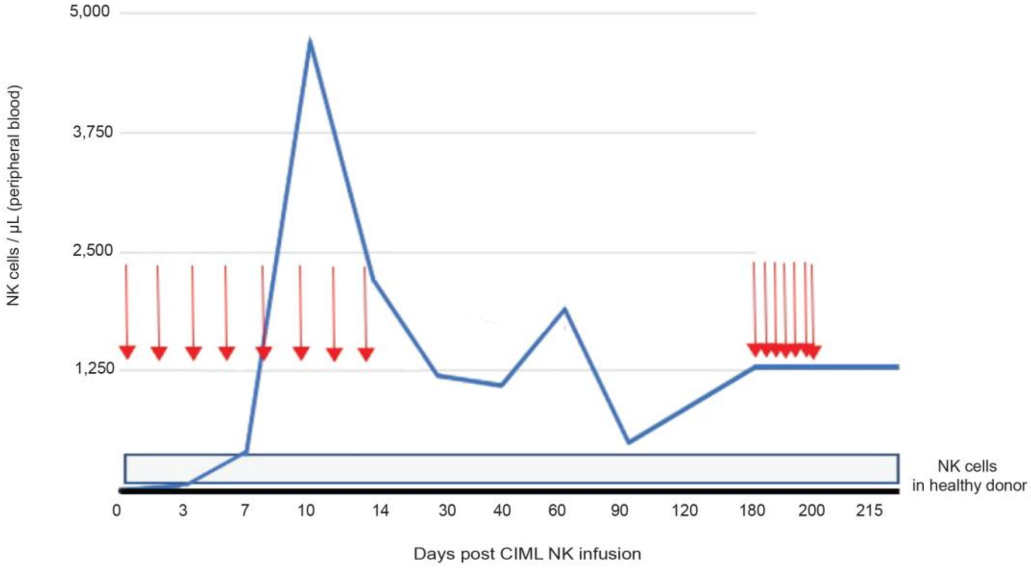


B

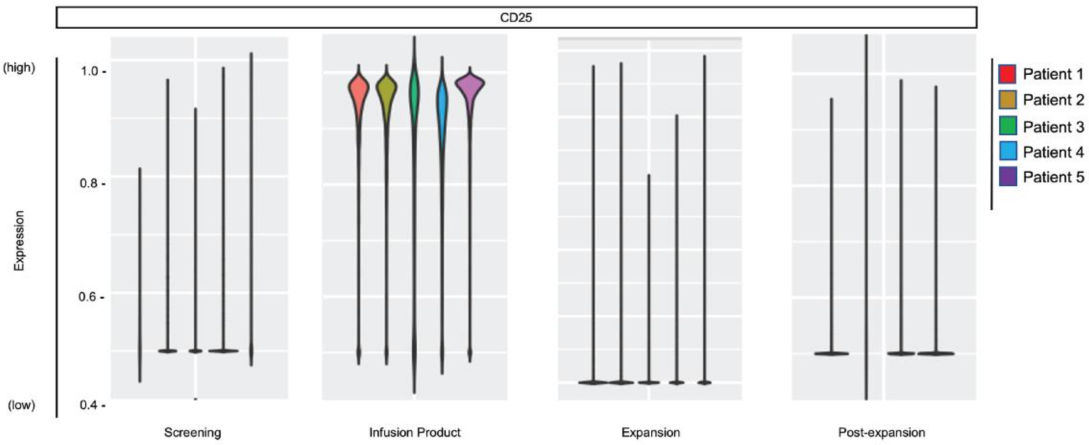


**Supplementary Figure S3.** CIML NK cell expansion mediated by low dose IL-2. **A** Total NK cell numbers per uL in the peripheral blood over time following infusion of CIML NK cells in Patient 1. Red arrows represent individual IL-2 injections. This patient received an additional set of IL-2 injections following relapse of disease in an attempt to reactivate the large amount of residual NK cells in the peripheral blood. There was no change with respect to total NK cell numbers following additional IL-2 injections. **B** Measurement of the IL-2 receptor CD25 on peripheral blood mononuclear cells at the indicated time points in each of the treated patients. Pre-infusion refers to the CIML NK products immediately before they were infused into patients, early expansion corresponds to days +7-14 after CIML NK infusion, late expansion corresponds to days +21-28 after CIML NK infusion, and post-expansion corresponds to day+60 or later after CIML NK infusion. Scaled expression of mass cytometry data is indicated on the y-axis, with 5 representing maximum expression and 0 no expression.

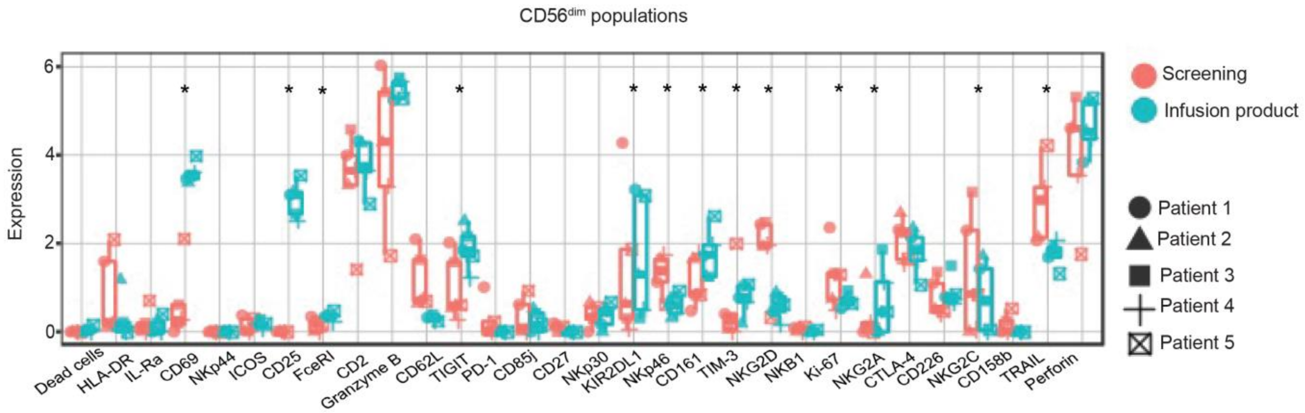


**Supplementary Figure S4**. Differential expression of CYTOF markers on NK cells between infusion and screening time points. $CD56^{dim}CD3^{-}$ NK cell clusters were identified in CyTOF marker expression data from PBMCs at both the screening and the CIML NK infusion product samples from all 5 patients. The expression of the markers indicated on the x-axis was compared between the NK cell clusters at the two timepoints. * p<0.05 by Wilcoxon rank sum test.

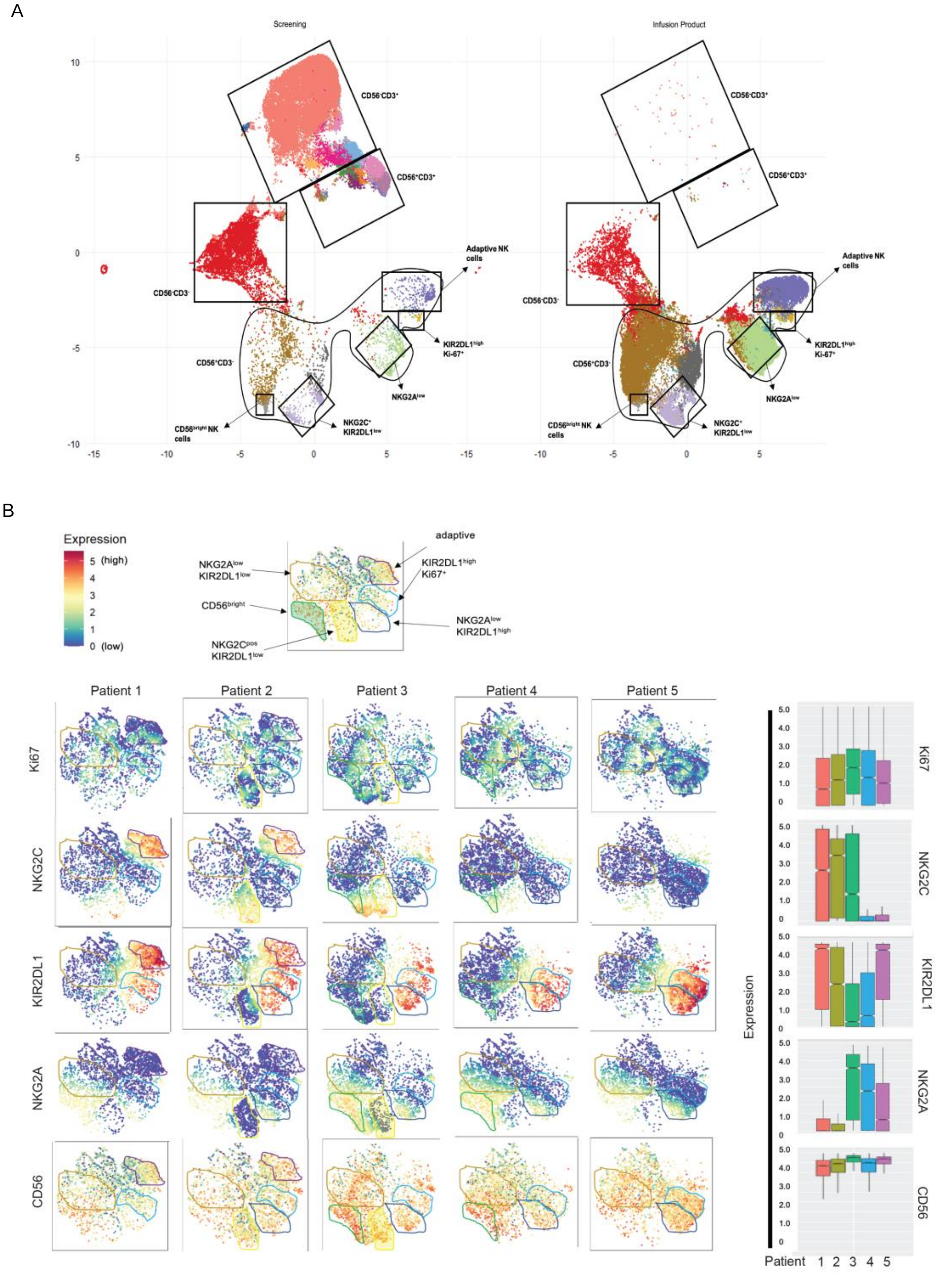


**Supplementary Figure 5.** Deep immunophenotypic analysis of the CIML NK infusion products in the 5 treated patients. **A** UMAP of the PBMCs at the screening time point and of the CIML NK infusion product. Data from all patients on the trial was aggregated and used for clustering. Specific populations are demarcated and labelled as shown. **B** tSNE plots corresponding to CIML NK products infused into individual patients is shown. Every column corresponds to a patient, and every row corresponds to a marker. Scaled expression of each marker is represented by the color scheme indicated in the legend, with a colorimetric gradient between high expression (red) and low expression (blue) shown at the top left. Total marker expressions plotted with notched boxplots comparing the 5 patients are shown on the right. Inter-patient heterogeneity in the expression of the indicated markers is noted in the expression of NKG2A, KIR2DL1 and NKG2C. Clusters corresponding to distinct NK cell subpopulations are indicated on all tSNE plots if they are present. The identity of these clusters is indicated in the representative tSNE plot at the top.

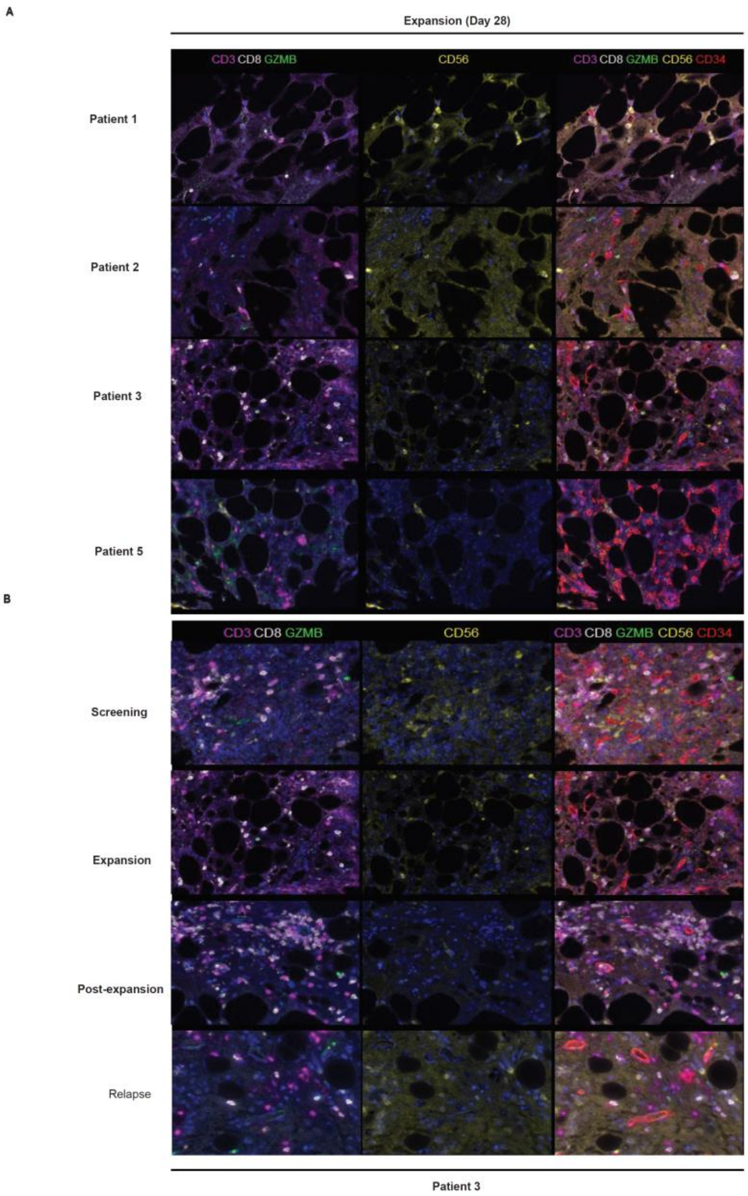


**Supplementary Figure S6.** Multiparameter immunofluorescence of patient bone marrow biopsies at the day+28 response assessment after CIML NK cell infusion. **A** The day+28 (expansion time point) after CIML NK cell infusion bone marrow biopsies are shown at the top, with each row corresponding to each of the patients who had bone marrow involvement of their disease at the time of infusion. The color corresponding to each marker is indicated at the top of each corresponding column, with juxtaposition of all markers shown in the rightmost column. $CD3^-CD56^+$ NK cells are present at the day+28 time point in all treated patients who had bone marrow involvement of their disease. **B** Longitudinal evaluation of indicated markers at the indicated time points in patient 3. A significant amount of $CD8^+$ cells are noted in the marrow at expansion and post-expansion time points. The time point labeled "persistence" refers to the post-expansion time point associated with overt relapse. $CD3^-CD56^+$ cells are largely not detectable in the bone marrow at post-expansion and persistence (relapse) time points.

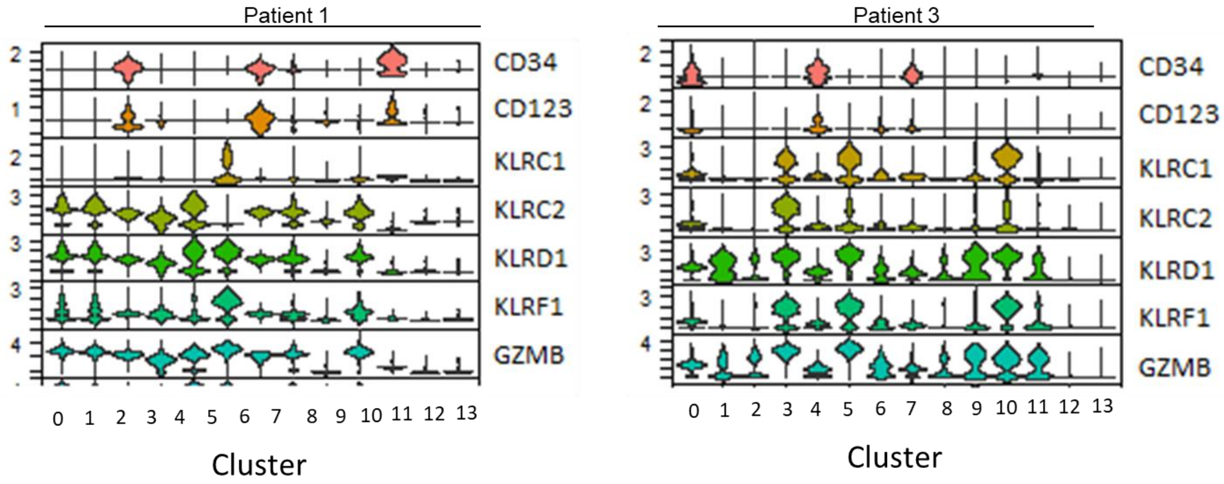


**Supplementary Figure S7.** Identification of disease and NK cell clusters using single-cell RNA sequencing at the time of relapse in Patients 1 & 3. Both patients' leukemia samples were positive for CD34 and CD123 at diagnosis.

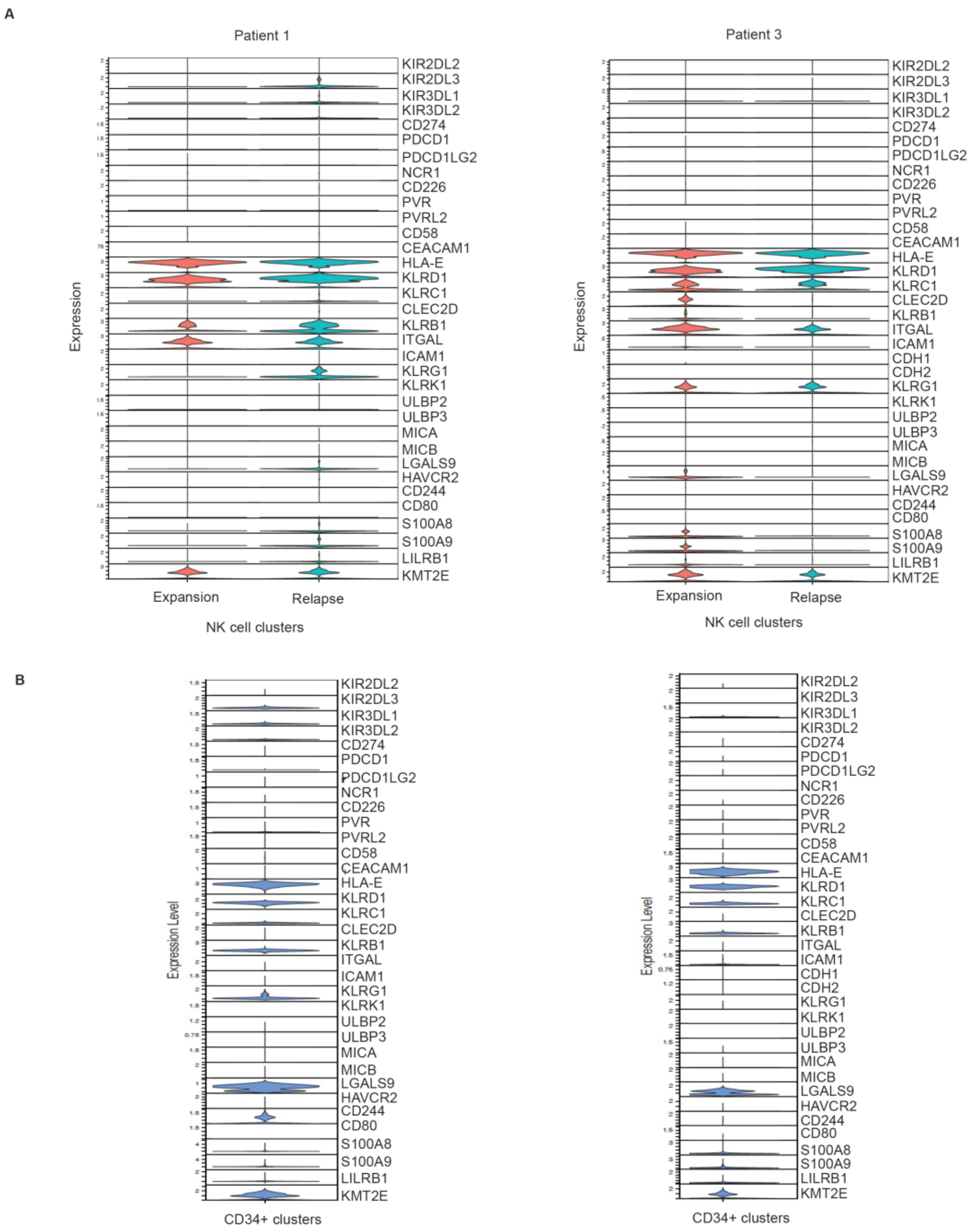


**Supplementary Figure S8.** Single-cell RNA sequencing of peripheral blood mononuclear cells from Patient 1 (left) and Patient 3 (right) at the Expansion and Relapse time points, with the cells from the latter fraction separated into CD34-depleted (CD34-) fractions (A) and CD34-enriched (CD34+) fractions (B). A Within the CD34-depleted fraction, NK cell clusters were identified based on the expression of the markers NCAM1, KLRD1, KLRF1, NCR1, NKG7, GNLY, CD3D, CD3E, and CD3G. Violin plots show gene expression of known NK receptors on the NK cell clusters in both patients. B Within the CD34-enriched fraction, clusters corresponding to leukemia blasts were identified based on the expression of the marker CD123 and absence of the aforementioned NK cell markers. Violin plots show the expression of known NK activating and inhibitory ligands on leukemia blasts in Patient 1 (left) and Patient 3 (right).
